# Circulating eNAMPT in Glaucoma: A Semi-Quantitative Plasma Analysis Before and After Nicotinamide Supplementation

**DOI:** 10.1101/2025.08.15.25333760

**Authors:** Antoni Vallbona-Garcia, Simon Gustavsson, Theo GMF Gorgels, James R Tribble, Carroll AB Webers, Hubert JM Smeets, Gauti Jóhannesson, Pete A Williams, Birke J Benedikter

## Abstract

Glaucoma is a neurodegenerative disease characterized by progressive retinal ganglion cell degeneration. Nicotinamide supplementation has demonstrated neuroprotective potential in glaucoma. Oral nicotinamide supplementation raises retinal and optic nerve levels of nicotinamide adenine dinucleotide (NAD) through the NAD salvage pathway, a process dependent on the rate-limiting enzyme nicotinamide phosphoribosyltransferase (NAMPT). Current evidence supports that NAMPT is essential for vision and retinal function, and its extracellular form (eNAMPT) has been detected in blood. Reduced levels of eNAMPT in blood could indicate impaired NAD biosynthetic capacity, and therefore, glaucomatous neurodegeneration susceptibility. This study aimed to (i) develop a specific, semi-quantitative assay to detect eNAMPT in plasma from glaucoma patients and controls, (ii) explore its potential as biomarker, and (iii) assess the effect of 2 weeks accelerated dosing nicotinamide supplementation on its circulating levels. This was done in samples from participants of a prospective clinical trial at the Eye Clinic, Umeå University Hospital (Sweden), which included 30 controls and 90 glaucoma patients that received oral 1.5 g/day nicotinamide for one week, followed by 3.0 g/day in the second week. A Western blotting assay was designed to detect eNAMPT (52 kDa) and transferrin (77 kDa) as housekeeping protein from 0.2 μL of EDTA plasma. Intra- and inter-assay variability of the assay were 14.9% and 37.9%, respectively. Normalized eNAMPT levels did not differ between glaucoma and controls, nor did they change following 2 weeks nicotinamide supplementation. In conclusion, eNAMPT is readily and specifically detected by Western blotting in EDTA plasma from controls and glaucoma patients. Given the role of NAD / NAMPT in neurodegenerative diseases, this study provides a platform for the specific detection of eNAMPT in liquid biopsies. Further studies specifically designed to study eNAMPT are needed to clarify its role in retinal ganglion cell degeneration and the therapeutic response to NAM.

## Introduction

Glaucoma is a group of neurodegenerative diseases hallmarked by the progressive dysfunction and degeneration of retinal ganglion cells (RGCs), the neurons that form the optic nerve. It is the leading cause of irreversible blindness, with more than 80 million people being affected worldwide. Elevated intraocular pressure (IOP) is the most prominent glaucoma risk factor. However, current treatments targeting IOP do not prevent disease progression in all cases, underlining the complex pathology of the disease.^1–3^ Since RGCs have high cellular energy requirements, it has been proposed that they might be particularly susceptible to bioenergetic impairments, and therefore, mitochondrial dysfunction.^4–7^ Novel therapies aiming to boost neuroprotection and survivability through improving the bioenergetic state of the RGCs are being explored, e.g., targeting NAD-related pathways which have progressed to Phase II and Phase III clinical trials.^8–10^

Nicotinamide (NAM), the amide form of vitamin B_3_ and a precursor of nicotinamide adenine dinucleotide (NAD), has demonstrated neuroprotective potential for the treatment of glaucoma, and clinical trials with its supplementation are currently ongoing (NCT05275738).^11–13^ NAM supplementation enhances NAD generation through the NAD salvage pathway.^14^ NAD’s primary form NAD^+^ plays essential roles in maintaining redox balance, supporting biosynthetic pathways, modulating cellular signaling, epigenetic modification, immunity, and DNA repair mechanisms.^14^ Its reduced form, NADH, serves as the primary electron donor for mitochondrial ATP production via the oxidative phosphorylation system (OXPHOS).^9,13–17^ Alterations in NAD metabolism have been associated with aging and age-related neurodegenerative diseases.^14,18^

The synthesis of NAD from NAM in the NAD salvage pathway depends on two enzymatic steps (Figure 1A). The first step depends on the rate-limiting enzyme nicotinamide phosphoribosyl transferase (NAMPT) to transform NAM into nicotinamide mononucleotide (NMN).^14,17^ The second step from NMN to NAD depends on the NMNAT1-2 enzymes. NAMPT-mediated NAD biosynthesis has been proposed to be essential for retinal function and vision in mice.^19^ In addition, inhibition of NAMPT enzymatic activity was shown to induce retinal toxicity.^20^ NAMPT is expressed in RGCs, albeit at lower levels than the second enzyme, of which the most predominant isoform in RGCs is NMNAT2.^21^ The expression of NAMPT has been shown to be reduced both in the RGCs of a glaucoma mouse model, as well as part of the optic nerve head (ONH) and the RGC layer of glaucoma patients, suggesting a decrease in the NAD salvage pathway capacity. Since RGCs highly favor NAD generation via the salvage pathway, a decrease salvage enzymes might be critical in disease.^11,21^

**Figure 1.**
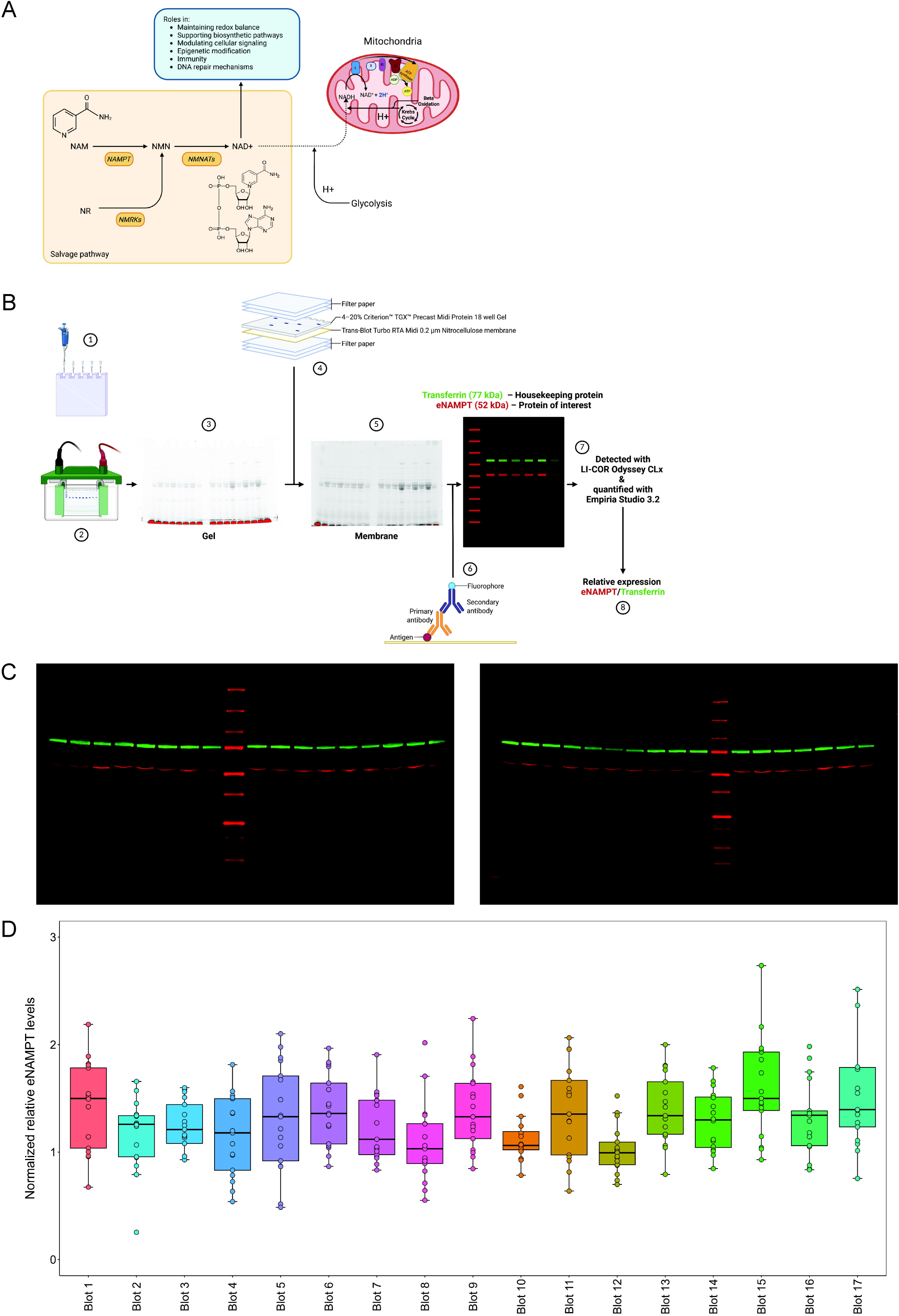
Summary of the WB assay to measure relative eNAMPT levels in plasma samples. A) Schematic overview of the NAD salvage pathway, highlighting various cellular roles, including its function as a mitochondrial electron donor in its reduced form (NADH). B) Schematic representation of the final WB workflow. In summary 1) 10 μL of plasma diluted 1:50 in Pierce Lane Marker Reducing Sample Buffer (1x) were loaded into a 4–20% Criterion TGX Stain-Free 18 well Protein Midi Gel and 2) run for 45 minutes at 200 V in a Criterion Cell. A reference sample is included in 3 positions in each gel for further blot corrections 3) Gel was imaged after electrophoretic separation with Geldoc Go Imaging System for the detection of the Stain-Free technology, allowing the visualization and confirmation of proper protein separation. 4) Gel separated proteins were transferred using the Trans-Blot Turbo Transfer System to a Midi 0.2 µm Nitrocellulose membrane and 5) visualization was again performed to confirm the proper transference and 1 hour blocking was performed at room temperature with slow agitation (∼40-45 rpm). 6) Primary antibody (eNAMPT protein of interest; Transferrin Housekeeping protein) overnight incubation following secondary antibody (IRDye 680RD; IRDye 800CW) 1 hour room temperature incubations were performed in black boxes with slow agitation (∼40-45 rpm). 7) Dynamic range detection of protein fluorescent bands was performed with the Li-Cor CLx imager and quantified with Empiria Studio 3.2 in order to 8) assess the relative eNAMPT expression to transferrin expression. C) Fluorescence images of 2 representative Western Blots. D) Distribution of the normalized relative eNAMPT plasma levels measured in all study cohort samples in each of the 17 performed WB. Normalized relative eNAMPT levels are calculated from the ratio of signals of eNAMPT by transferrin as housekeeping protein and corrected inter-blot wise by the reference sample values. eNAMPT: extracellular nicotinamide phosphoribosyltransferase; NAD: nicotinamide adenine dinucleotide; NAM: nicotinamide; NAMPT: nicotinamide phosphoribosyltransferase; NMN: nicotinamide mononucleotide; NMNATs: nicotinamide-nucleotide adenylyltransferases; NMRKs: nicotinamide riboside kinases NR: nicotinamide riboside; WB: Western blot.

NAMPT has also been detected in the bloodstream in an extracellular form; eNAMPT (alternatively named Visfatin or pre-B cell colony enhancing factor [PBEF]).^14,22,23^ eNAMPT differs from its intracellular form due to post-translational modifications and can be secreted by different cells e.g., cardiac cells, β-cells and adipocytes via an unknown unconventional pathway. This extracellular form is suggested to have a function as a cytokine.^22,24–29^ However, it has also been suggested that eNAMPT secreted in the bloodstream and encapsulated inside extracellular vesicles (EVs) is able to influence intracellular NAD biosynthesis in various cells types and tissues.^23,26,30,31^ In a mouse model with specific adipose tissue NAMPT overexpression, an increase in NAD has been observed in multiple tissues, including the retina.^23^

Given the possible importance of circulating eNAMPT levels in modulating NAD biosynthesis throughout the body, and its potential relationship with the retina, systemic eNAMPT levels might reflect the bioenergetic capacity of high-energy-demanding RGCs. Previous studies have reported systemic metabolic defects related to NAD metabolism in glaucoma.^32,33^ Likewise, levels of eNAMPT in blood may be impaired and could indicate metabolic vulnerability. Thus, levels of circulating eNAMPT may serve as biomarkers for glaucomatous neurodegeneration susceptibility. With current clinical trials ongoing on NAM supplementation in glaucoma with, it is important to study the systemic effects of this supplementation. NAM supplementation aims to increase cellular NAD levels, which in turn activate sirtuins such as SIRT1, whose deacetylation capacity has been proposed to enhance eNAMPT secretion and enzymatic activity, suggesting relevant mechanisms related to NAM therapy.^22,26,29,34–36^

Although commercial immunoassays such as ELISAs are available to detect circulating NAMPT, substantial variability in specificity and signal interpretation has been reported. Previous studies have shown inconsistent results across assay types and observed discrepancies in detected molecular forms of eNAMPT, highlighting the need for more specific detection methods.^37^

In the current study, we aimed to:

1. Establish a semi-quantitative and specific method to measure eNAMPT levels using Western blot (WB).
2. Investigate whether baseline eNAMPT levels differ between different types of glaucoma and healthy controls.
3. Explore whether 2 week oral NAM supplementation alters circulating eNAMPT levels and whether eNAMPT levels are associated with patient phenotypes.

In this study, we demonstrated that eNAMPT is readily available in the blood and can be detected with high specificity and sensitivity in glaucoma patients and control subjects.

## Materials and methods

### Study cohort, ethical approval and patient sampling

The cohort has been described in Gustavsson et al., 2023.^38^ In summary, a prospective clinical trial was conducted at the Eye Clinic at Umeå University Hospital, (Umeå, Sweden) which included 30 high tension glaucoma (HTG), 30 normal tension glaucoma (NTG), 30 pseudoexfoliative glaucoma (PEXG) and 30 age and sex-matched controls. During this prospective clinical trial, patients underwent a 2-week accelerated dosing NAM supplementation (1.5 g/day during the first week; 3g/day during the second week). Pre and post-treatment examinations were performed, together with collection of blood. The human studies adhered to the tenets of the Declaration of Helsinki, and the ethics protocols were approved by the Swedish Ethical Review Authority (2020-01525, 2021-01036, 2021-03745, and 2022-04851).

Venous blood samples were collected from each study participant in EDTA tubes at baseline and after NAM treatment, at least three hours after the last meal. The tubes were sent immediately after sampling to a centralized biobank (Biobanken Norr) in the same hospital where the samples were collected (University Hospital of Umea). At the biobank, the tubes underwent centrifugation (1500G, 15 minutes, room temperature), and aliquots (all 250 µL) were collected from the tubes using a Hamilton Microlab Star LET. Aliquots of erythrocytes, plasma, and buffy coat were collected from the EDTA tubes. All the aliquots were placed in SBS format cryotubes and then immediately stored at -80°C until further analysis. The time between sampling and storage was 37 – 180 minutes (mean value 80 minutes, median value 77 minutes). The aliquots were kept frozen at -80°C as they were sent for analysis. Out of the 120 participants included in the study, the following plasma samples were available and used in this study: 236 plasma samples (n = 30 controls, n = 30 HTG, n = 30 NTG, n = 29 PEXG; 119 pre-NAM / n = 30 controls, n = 29 HTG, n = 29 NTG, n = 29 PEXG; 117 post-NAM).

Some samples were not available due to the participant not being able to join the first or second study visit, or the blood fraction could not be properly collected and stored. Patient data and experimental data from this study can be found in Supplementary Data 1.

### Immunoblotting experimental optimization and design

Optimization of several immunoblotting aspects was performed, which are summarized in **Table 1**, such as the use of plasma or serum, the sample boiling time, volume of diluted sample to load, antibody concentrations, gel type, incubation protocol and normalization method. The procedures used for sample preparation and immunoblot execution are described in detail in the following sub-sections.

**Table 1.** Summary of the several parameters that were assessed for the optimization of the assay on the detection of extracellular nicotinamide phosphoribosyltransferase (eNAMPT) protein levels in human plasma. The parameters are listed chronologically in the order of testing, and following parameters were tested after fixing the parameters listed above.

| Variable tested | Conditions compared | Replicates | Key observation / conclusions | Condition chosen |
| --- | --- | --- | --- | --- |
| Sample material | EDTA plasma vs. serum | 3 donors | Similar bands signal and morphology | EDTA plasma because eNAMPT was reported in extracellular vesicles (EVs) and serum may contain >90% platelet-derived EVs |
| Sample heating to 95°C | 5 min vs. 30min | 3 donors | While Yoshida et al.<sup>23</sup> previously reported heating to 95°C for 30 min as requirement for clear eNAMPT bands, eNAMPT signal and band appearance was similar after 5 and 30 min in our hands. Transferrin shows a clear band after 5 minutes, but smaller and multiple bands at 30 min, reflecting possible degradation products. | 5 min at 95°C |
| Sample buffer | Pierce Reducing Lane Marker vs. Reducing Laemmli Buffer | 3 donors | Similar signal | Pierce Reducing Lane Marker |
| Volume of 1:50 diluted plasma | 25, 10, 5 and 2,5 µl | Reference sample | 10 µl gives clear bands for both eNAMPT and Transferrin without any evidence for fluorescence self-quenching. Fluorescence self-quenching is seen when loading 25 µl, while bands become incomplete for 5 and 2,5 µl. The signal increases with increasing loaded volume. | 10 µl |
| Dilution NAMPT antibody | 1:2500, 1:1000, 1:500 | Reference sample | 1:1000 shows a clear, complete, not self-quenched band. Band at 1:2500 and 1:500 show lower and higher signal respectively than 1:1000. | 1:1000 |
| Dilution transferrin antibody | 1:10000, 1:5000, 1:2500, 1:1000 | Reference sample | 1:5000 shows a clear, complete, not self-quenched band. 1:10000 shows a lower signal than 1:5000. 1:2500 and 1:1000 show a higher signal than 1:5000. | 1:5000 |
| Gel type | Tris-Glycine extended (Midi, 4-12%, 18 wells, Bio-Rad) / Bolt Bis-Tris (Mini 4-12% 17 wells, Thermo Fisher Scientific) | Reference sample | TGX Midi gels displayed clearer and flatter bands without smear for both proteins while Bolt Bis-Tris showed a strong smile. Better intra-assay variability was also observed on TGX Midi gels across the whole blot (sides and center). | Tris-Glycine extended (Midi, 4-12%, 18 wells) |
| Antibody incubation protocols | Type of shaking (rolling or flat) and volume of Ab/blocking (10 ml or 20 ml). | Reference sample | Flat incubation, shaking to all sides at ~40 rpm, with 20 ml of antibody solution in a closed black box gave the most stable less intra-assay variable results on both bands. Rolling incubation and lower volume were linked with higher intra-blot variability / staining gradients. | Flat incubation with 20 ml |
| Normalization measurement | Total protein vs. transferrin | Reference sample | We explored the total protein staining (Stain-Free technology) on the imaged membranes as an alternative method to transferrin as housekeeping normalization protein (See methods). Quantified total protein did not correlate to the band signals. Possible explanations are (1) high background on total protein images (membrane wet due to transfer buffer) and (2) imaging on a different device (GelDoc Go, Bio-Rad) than fluorescent antibodies (Odyssey CLx, Li-Cor). | Transferrin as housekeeping protein to normalize eNAMPT |

With the optimized protocol, a total of 17 Western blots were performed to analyze all 236 plasma samples of the study cohort. Each blot contains samples from 6-7 randomized study participants, with the respective treatment paired sample of the subject in the same blot adjacent to it. In all the blots, the same plasma reference sample was situated in 3 different positions (both extreme sides and center of the gel) in order to assess the intra and inter-blot variability, and as a correction/normalization factor allowing inter-blot standardization.

### Plasma sample preparation for immunoblot

Plasma aliquots stored at -80°C were altogether thawed O/N on ice and inside a cold room (4°C). Plasma was diluted 1:50 in Pierce™ Lane Marker Reducing Sample Buffer (Thermofisher, 39000; diluted stock from 5X to 1X in Milli-Q water). Samples were warmed at 95°C for 5 minutes, cooled immediately back on ice, and then frozen at -80°C.

### Immunoblotting

Per blot, plasma samples in sample buffer were thawed for 10 minutes at RT. Then samples were spun down and vortexed prior to loading 10 ul in 4–20% Criterion™ TGX Stain-Free™ 18 well Protein Gel (Bio-Rad, #5678094). 2 ul of Odyssey One-Color Protein Molecular Weight Marker (Li-Cor, 928-40000), for visible viewing and 700 nm channel near-infrared detection, was also loaded in each gel. Electrophoretic separations were performed at 200 V for 45 minutes using running buffer containing 25 mM Tris, 192 mM glycine, 0.1% SDS and pH 8.3, in the Criterion Cell (Bio-Rad, 1656001). After electrophoretic separation, gels were imaged with the Geldoc Go Imaging System (Bio-Rad, #12009077) using the Stain-Free Gel mode (45 sec activation, 1-3 sec exposure) for activation of the Stain-Free technology, confirming the proper protein separation. Then, the proteins were blotted using the Trans-Blot Turbo RTA Midi 0.2 µm Nitrocellulose Transfer Kit (#1704271), according to manufacturer instructions, and the Trans-Blot® Turbo™ Transfer System (Bio-Rad #1704150) with the following MIDI protocol settings: 2.5 A and 25 V for 7 minutes. The blot after transference was imaged with the Geldoc Go Imaging System (Bio-Rad, #12009077) using the Stain-Free Blot mode (1-3 seconds exposure) to confirm the proper protein transfer.

The blot was blocked for 1 hour at room temperature in 20 ml of Intercept® PBS Blocking Buffer (Li-Cor, 927-70003) diluted 1:2 in PBS 1X (w/o Ca, w/o Mg), with slow agitation in a plate shaker (∼40-45 rpm). After that, 3 washes of 5 minutes each with freshly prepared PBS 1X (w/o Ca, w/o Mg) with 0.1% Tween were performed. Then an overnight incubation at 4°C with slow agitation in a plate shaker (∼40-45 rpm) was performed with 20 ml of Intercept® PBS Blocking Buffer (Li-Cor, 927-70001) diluted 1:2 in PBS 1X (w/o Ca, w/o Mg), containing primary antibodies with the following dilutions: 1:1000 NAMPT (Adipogen, AG-20A-0034-C100, mouse monoclonal clone OMNI-379) and 1:5000 Transferrin (Abcam, ab82411, rabbit polyclonal).

After the incubation with the primary antibody, membranes were washed 3 times for 5 minutes each with freshly prepared PBS 1X (w/o Ca, w/o Mg) with 0.1% Tween. Membranes were then incubated at room temperature for 1 hour in 20 ml of Intercept® PBS Blocking Buffer (Li-Cor, 927-70001) diluted 1:2 in PBS 1X (w/o Ca, w/o Mg) containing goat anti-mouse IRDye 680RD (1:10000, 926-68070) and goat anti rabbit IRDye 800CW (1:10000, 926-32211). Membranes were washed 3 times for 5 minutes each with freshly prepared PBS 1X (w/o Ca, w/o Mg) with 0.1% Tween and dried 1 hour in filter paper prior to imaging.

### Blot imaging

Dried membranes were imaged with the Li-Cor CLx imager with the following settings: offset 0.5 mm, lowest quality, scan resolution 169 μm, and autoexposure for both the 700 and the 800 channels. This allowed the detection of both fluorescent antibodies, and of the Odyssey One-Color Protein Molecular Weight Marker in the 700 nm channel.

### Image analysis

Image analysis was performed with Empiria Studio 3.2 in order to quantify both eNAMPT (protein of interest) and transferrin (housekeeping protein) bands per plasma sample. This is done using the Housekeeping and Targets analysis mode and automatic band detection of the mentioned software.

Quantification of Stain-Free total protein images (.tiff) obtained with the Geldoc Go Imaging System (Bio-Rad, #12009077) after imaging the transferred membrane was performed using ImageJ 1.54f with an inhouse macro. The mean intensity of the pixels of the portion of bands between 30 and 100 kDa per plasma sample was quantified. The macro allowed the adjustment of the TIF Stain-Free image to the right angle for the proper quantification and allowed to use the same size box for the quantification of protein and background levels between blots.

### Analysis and statistics

The normalized relative eNAMPT levels per subject were analyzed using generalized linear mixed-effects modeling (Gamma distribution with a log link and Gaussian random effects), including in the fixed effects the study groups (HTG, NTG, PEXG or Control), NAM treatment effect, and the interaction between the treatment and the groups. The model also contained adjustments for phenotypic covariates (sex, age). In the random effects, the variable ‘Subject’ was included in order to pair subjects before and after treatment.

The inference criterion used for comparing models is their ability to predict the observed data, i.e., models are compared directly through their minimized minus log-likelihood. When the numbers of parameters in models differ, they are penalized by adding the number of estimated parameters, a form of Akaike information criterion (AIC).^39^ Using the AIC, we found that a generalized linear mixed-effects model with a Gamma distribution with a Log link and Gaussian random effects fit the normalized relative eNAMPT levels better than a linear model and generalized linear models with other distributions and links.

Possible relations of normalized relative eNAMPT levels in each study group prior to and post-NAM treatment were assessed through Pearson correlations with the disease severity measured through visual field index (%), and optical coherence tomography angiography (OCTA) values from the ONH and the macula. All statistical analysis presented in regard to the normalized relative eNAMPT levels were performed using R v4.3.1, and the ‘glmer’ function of the publicly available library ‘lme4’.^40,41^

## Results

### Validation of semi-quantitative eNAMPT detection and assay variability

A semi quantitative fluorescent WB assay was designed to detect and quantify the circulating eNAMPT protein levels. Details on the assay optimization are summarized in Table 1. In the final assay setup, 236 human EDTA plasma samples were analyzed on 17 WBs (workflow illustrated in Figure 1B). Each WB included the same reference plasma sample in 3 different positions to monitor intra- and inter-blot variability, as well as matched pre- and post-NAM supplementation samples from 6-7 randomized study participants from different glaucoma (HTG, NTG, PEXG) and control groups (see study cohort in the results section). A compilation of all raw WB can be found in Supplementary Figure 1.

eNAMPT was detected in all samples with a single specific band at 52 kDa (Figure 1C, Supplementary Figure 1), using only 0.2 μL of plasma (10 µl of 1:50 dilution). The abundant plasma protein transferrin was used as housekeeping protein, as common intracellular housekeeping proteins are not suitable for this cell-free sample. Transferrin was also detected in all plasma samples at the same dilution with a band at 77 kDa (Figure 1C, Supplementary Figure 1).

Using the reference sample, we analyzed the intra- and inter-assay variability for raw eNAMPT and transferrin signals, as well as the eNAMPT/transferrin ratio. For the eNAMPT/transferrin ratio, the intra-assay CV was of 14.9 ± 8.4 % and the inter-assay CV was of 37.9%. Detailed information on variability can be found in Supplementary Table 1 and Supplementary Figure 2.

Due to the high inter-assay CV, normalization was performed by dividing each sample’s eNAMPT/transferrin ratio by the average eNAMPT/transferrin ratio of the reference sample on the respective blot. The normalized values are referred to as “normalized relative eNAMPT levels”, and these normalized values were used for the analysis of the study cohort. The normalized values of all samples in the study in each WB assay are shown in Figure 1D.

### Study cohort

The cohort has been described in Gustavsson et al., 2023.^38^ Patient data and experimental data from this study can be found in Supplementary Data 1.

### Systemic eNAMPT protein levels pre and post NAM treatment

After quantifying relative eNAMPT plasma levels according to the optimized workflow described above, we moved on to study baseline differences in eNAMPT between the glaucoma groups (HTG, NTG, PEXG) and healthy controls. We also analyzed the possible within-group effect of the 2-week oral NAM treatment on eNAMPT. The analysis was performed through a paired generalized linear mixed model with study groups, treatment effect, and their interaction (See methods). The model also included covariate adjustment for age and sex (Supplementary Table 2).

We did not observe significant differences in eNAMPT levels between the glaucoma groups and healthy controls at baseline (Figure 2A). After NAM treatment, eNAMPT levels also remained similar between the groups (Figure 2B). Regarding a possible effect of oral NAM treatment on eNAMPT levels, we did not observe a significant change within the study groups (Figure 2C). In relation to covariates, no significant relationship was observed between eNAMPT levels and age or sex, either generally or within specific study groups (Figure 3A, 3B). However, within sexes, we observed that females from all three glaucoma groups presented a tendency for lower eNAMPT levels in comparison to female controls (Figure 3B). This is similarly observed at baseline and after NAM treatment, and it is not seen in males. Finally, we did not observe a relation of eNAMPT levels with glaucoma severity measured through visual field index (%) (Figure 3C).

**Figure 2.**
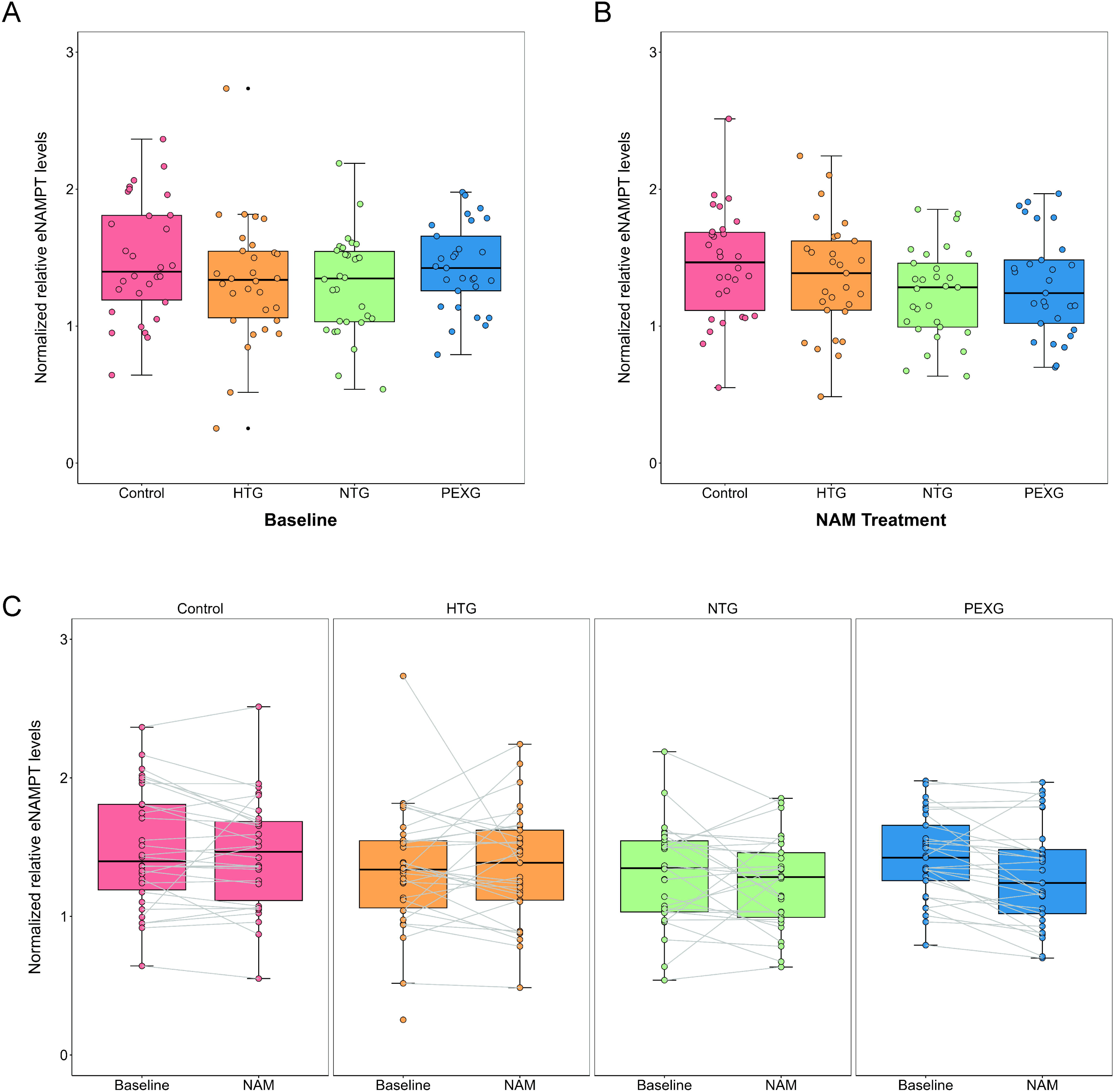
Normalized relative eNAMPT levels in plasma are comparable between glaucoma subtypes and the control group, and no within-group treatment effect is observed following oral NAM treatment. A, B) Analysis through a paired generalized linear mixed-model with covariates (age, sex) showed no differences in the normalized relative eNAMPT plasma levels between glaucoma subtypes and controls at baseline (pre-NAM treatment) (A) or after NAM treatment (post-NAM treatment) (B). C) No significant within-group effect of NAM treatment on normalized relative eNAMPT levels in plasma was observed in the paired generalized linear mixed-model with covariates (age, sex). Normalized relative eNAMPT levels are calculated from the ratio of signals of eNAMPT by transferrin as housekeeping protein and corrected inter-blot wise by the reference sample values. HTG: high tension glaucoma; NAM: Nicotinamide; eNAMPT: extracellular nicotinamide phosphoribosyltransferase; NTG: normal tension glaucoma; PEXG: pseudoexfoliative glaucoma.

**Figure 3.**
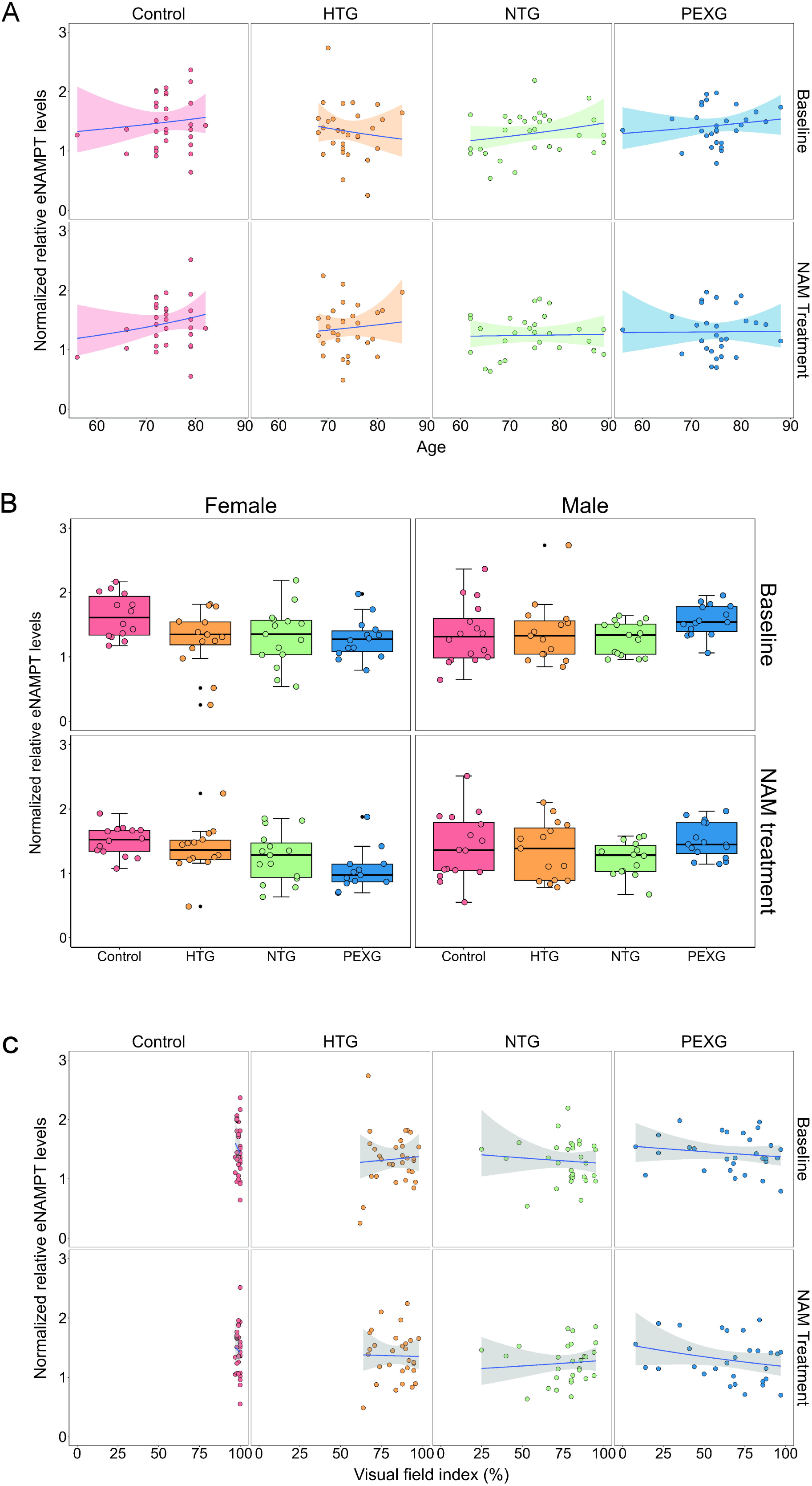
Normalized relative eNAMPT levels in plasma and its relation with covariates age and sex, and with glaucoma severity measured through visual field index (%). A) Analysis through a paired generalized linear mixed-model with covariates (age, sex) showed no relation of the normalized relative eNAMPT plasma levels with age in general nor in specific groups, at baseline (pre-NAM treatment) or after NAM treatment (post-NAM treatment). B) No significant relation between normalized relative eNAMPT plasma levels and sex in the mentioned mixed-model was observed, although a tendency for lower levels in glaucoma females than control females was observed. C) No relation between relative eNAMPT levels and glaucoma severity measured by visual field index (%) pre- and post-NAM treatment was observed. Normalized relative eNAMPT levels are calculated from the ratio of signals of eNAMPT by transferrin as housekeeping protein and corrected inter-blot wise by the reference sample values. HTG: high tension glaucoma; NAM: Nicotinamide; eNAMPT: extracellular nicotinamide phosphoribosyltransferase; NTG: normal tension glaucoma; PEXG: pseudoexfoliative glaucoma.

### eNAMPT levels do not correlate with OCTA parameters

We also investigated the potential relationship between eNAMPT levels and retinal vascular parameters measured by OCTA in either the ONH or the macula. However, no association was observed between eNAMPT levels and OCTA values in either glaucoma patients or controls, at baseline or following NAM treatment (Supplementary Figures 3 and 4).

## Discussion

In the current study, we established a semi-quantitative, specific and sensitive method to detect eNAMPT in plasma and investigate its levels pre- and post-NAM supplementations in glaucoma and corresponding controls. NAMPT is the rate-limiting enzyme of the NAD salvage biosynthetic pathway, which has been proposed to be crucial for retinal function. Circulating blood eNAMPT levels have been proposed to be able to modulate NAD biosynthesis throughout the body, including the retina. Therefore, systemic eNAMPT levels might reflect the bioenergetic capacity of high-energy-demanding cells, such as RGCs, with impaired levels indicating defects in the NAD biosynthetic salvage pathway and thus serving as a marker of metabolic vulnerability. Treatments, such as NAM supplementation, that boost NAD biosynthesis through the NAD salvage pathway are arising as complementary therapies in glaucoma, aiming to improve RGC bioenergetics. Thus, it is also important to study the effects of NAM supplementation on NAD biosynthetic pathways.

### Technical aspects of the eNAMPT detection by WB

We have used WB to analyze eNAMPT in EDTA plasma, while earlier eNAMPT studies have generally used ELISA either performed in serum or plasma.^42–47^ The reason for choosing WB is that inconsistent results have been reported in a direct comparison of different immunoassay types, probably due to discrepancies in the detection of molecular forms of eNAMPT.^37^ WB offers a higher specificity for eNAMPT detection by enabling the detection of the protein of interest at the correct molecular weight. However, WB technical aspects, e.g., protein electrophoresis, protein transfer to a membrane, blocking, antibody binding, and quantification of fluorescence signals from protein bands, lead to a rather high technical variability. This variability was reduced during protocol optimization, and a threshold of 15% intra-blot CV was reached across several WB performed. While WB allows specific protein detection by size, the throughput is limited. This led to the use of single technical replicates in order to assess the 236 plasma samples in a total of 17 WB assays.

### Association between eNAMPT and glaucoma

The amount of eNAMPT in plasma was similar between controls and the 3 glaucoma subtypes, HTG, NTG and PEXG. This suggests that if bloodstream eNAMPT levels influence the intracellular salvage pathway capacity to synthesize NAD in retinal tissue, this would not be compromised in glaucoma. Alternatively, our assay might lack the resolution to detect subtle quantitative differences with the current sample size, as the study was not powered for the assay’s variability.

With regard to phenotypic traits, no relation of eNAMPT with age was observed. Previous studies showed a decline with age in humans, which might be explained by the age range including younger donors (∼40 to 80 years old) in comparison to those in this study (56 to 89 years old).^23^ With regard to sex, no relation was observed with eNAMPT levels. However, although not statistically significant, females from all 3 glaucoma subtypes tended to exhibit lower eNAMPT levels compared to female controls. Previous studies that measured eNAMPT levels in serum through ELISA have reported sex differences. In these studies, diurnal variation of eNAMPT levels and a correlation with body weight were also observed.^48,49^ A possible decrease of eNAMPT levels in females with glaucoma could be studied further, taking into account these potential confounders.

While this was the first study to investigate eNAMPT in the context of glaucoma, Kaja et al.^42^ previously assessed eNAMPT as a biomarker in subjects with a history of retinal vascular occlusion, although with a different sample type and method. In these subjects, reduced serum eNAMPT levels, measured through ELISA, were observed. The authors suggested that low bloodstream levels of eNAMPT might indicate low intracellular levels and activity, leading to a reduced endogenous protection against ischemic events.^42^ Vascular pathological situations are also suggested to happen in glaucoma. The vascular theory in glaucoma proposes that diminished perfusion pressure, impaired vascular autoregulation, and disrupted neurovascular coupling contribute to the RGC and, therefore, optic nerve degeneration.^50,51^ In the current glaucoma cohort, Gustavsson et al.^38^ previously observed impaired retinal vasculature parameters in the ONH and macula in glaucoma patients in comparison to controls. However, we found no correlation between eNAMPT and these retinal vascular outcomes. As mentioned, our results suggest that if eNAMPT levels in blood can influence retinal capacity to synthesize NAD, this would not be affected in glaucoma. The NAD salvage pathway enzymatic machinery has been observed to be completely expressed in single-cell and single-nucleus RNA-seq datasets from post-mortem retinas, specifically in RGCs.^21^ This shows the capacity of the cells to independently execute this pathway, in line with a potentially limited importance of eNAMPT for the bioenergetic status of RGCs. While NAMPT has been shown to be reduced in glaucoma retinal tissue from late-stage patients in the same study, this might only suggest the loss of neuronal tissue in the retina and ONH due to severe disease.

### Mechanisms of eNAMPT secretion and its function

In animal studies, increased eNAMPT in the bloodstream has been observed to correlate with an increase in NAD levels in organs such as the hypothalamus or the retina.^23,48^ The mechanism of how eNAMPT is extracellularly secreted, is internalized by cells, and is able to influence their salvage pathway capacity is not fully known. However, it is suggested to occur only with NAMPT that is encapsulated and delivered in EVs.^23,30^ eNAMPT was first described as adipokine named Visfatin/pre–B cell colony-enhancing factor (PBEF), having diverse functions in tissues and disease, and being secreted by different cells such as adipocytes or cardiac cells.^24–28^ The current study assessed total eNAMPT, as WB sample buffer contains 5% SDS, breaking those EVs and thereby assessing proteins outside and within EVs.^52^ In the future, it could be further investigated whether EV-encapsulated eNAMPT has distinct biomarker potential and functional activity in the context of glaucoma, as compared to unencapsulated eNAMPT.

### Effect of NAM supplementation on eNAMPT

NAM supplementation aims to increase NAD production. The NAD formed performs critical functions at different levels in the cell.^14^ Increased NAD^+^ levels have been shown to activate sirtuins, NAD^+^-dependent histone deacetylases (HDACs), and specifically SIRT1. SIRT1 can directly interact and influence the activation of PGC1-α, a master regulator of mitochondrial biogenesis.^34–36,53,54^ Interestingly, SIRT1 has been observed to deacetylate NAMPT in its lysine 53, which is suggested to facilitate its release as well as increase the activity of the enzyme.^22,26,29^ Therefore, NAM supplementation could lead to an increase in NAD pools, activating SIRT1 and, therefore, promoting the secretion of NAMPT. However, administering NAM supplements orally for 2 weeks did not lead to a change in circulating eNAMPT. Main sources of eNAMPT in the bloodstream are suggested to be cardiac cells, β-cells and adipocytes.^24–28^ The mentioned cells are not disease-affected in glaucoma, and their status might be optimal with eNAMPT secretion not being further enhanced by an increase in NAD.

## Limitations

The relatively high variability of the WB technique may interfere with detection of mild inter-group differences. Secondly, the technique does not distinguish two (functionally) distinct forms of eNAMPT, within and outside of EVs. Thirdly, the clinical study was not initially designed for our research aims. The duration of NAM supplementation was short and the limited sample size per subgroup may have prevented the detection of differences related to phenotypic traits e.g., for instance sex-specific differences in eNAMPT between glaucoma patients and controls.

## Conclusion

In conclusion, eNAMPT is readily and specifically detected by Western blotting in EDTA plasma from controls and glaucoma patients. Given the role of NAD / NAMPT in neurodegenerative diseases, this study provides a platform for the specific detection of eNAMPT in liquid biopsies. Further studies specifically designed to study eNAMPT are needed to clarify its role in RGC degeneration and the therapeutic response to NAM.

## Data Availability

All data generated or analyzed during this study will be included in the final peer-reviewed published article.

## Declarations

## Ethics approval

The study was approved by the Swedish Ethical Review Authority (2020-01525, 2021-01036, 2021-03745, and 2022-04851).

## Consent for publication

Not applicable.

## Author’s contributions

AVG –conceived and designed experiments, performed experiments, analyzed and interpreted data, wrote the manuscript.

SG – conceived and designed experiments, performed experiments, analyzed and interpreted data, wrote the manuscript.

TGMF – conceived and designed experiments, provided supervision, wrote the manuscript.

JRT – provided supervision, wrote the manuscript.

CAB – provided supervision, wrote the manuscript.

HJM – provided supervision, wrote the manuscript.

GJ – provided supervision, conceived and designed experiments, wrote the manuscript.

PAW – provided supervision, conceived and designed experiments, wrote the manuscript.

BJB – acquired funding, provided supervision, conceived and designed experiments, interpreted data, wrote the manuscript.

All authors read and approved the final manuscript.

## Acknowledgements

The authors would like to thank St. Erik Eye Hospital for financial support for research space and facilities. The authors would like to thank the Dutch Eye Foundation, which contributed through UitZicht.

