## Supplementary figure 1 for "Circulating eNAMPT in Glaucoma: A Semi-Quantitative Plasma Analysis Before and After Nicotinamide Supplementation"

**Supplementary figure 1. Compilation of all Western Blots.** Includes total protein signal on each of the 17 gels after electrophoresis or membranes after protein transfer, as well as fluorescent antibody staining for transferrin (77 kDa, green) and NAMPT (52 kDa, red). The reference sample positions are marked with a red *. Ab: Antibody; eNAMPT: extracellular nicotinamide phosphoribosyltransferase.

1 Gel Membrane Ab staining

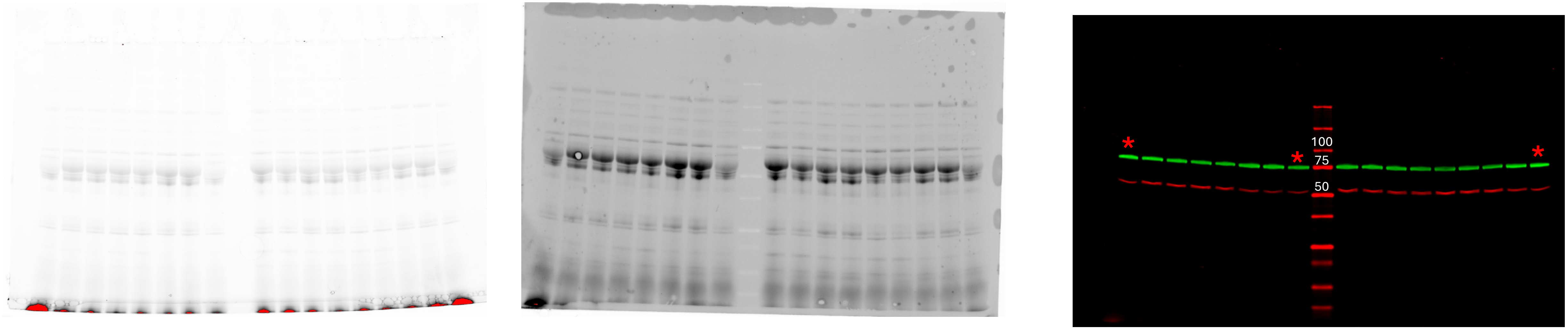


2

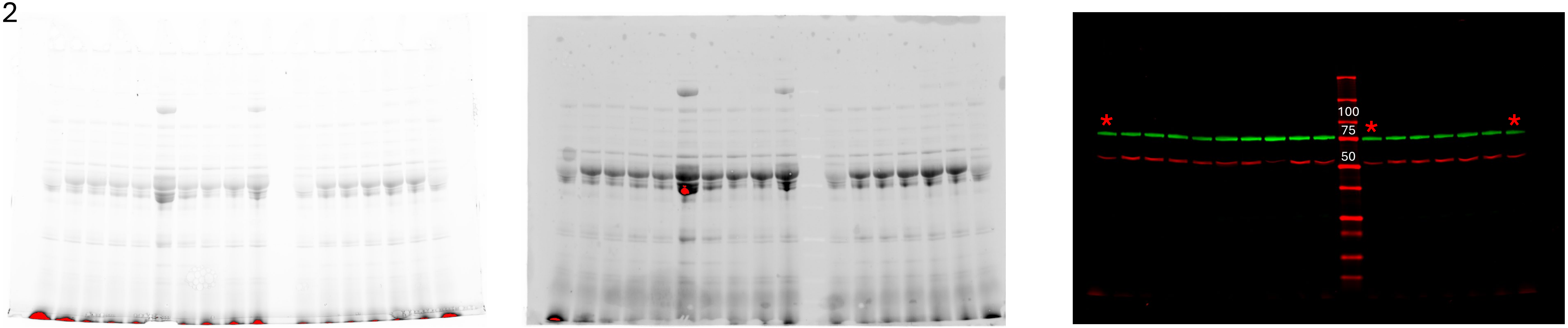

3 Gel Membrane Ab staining

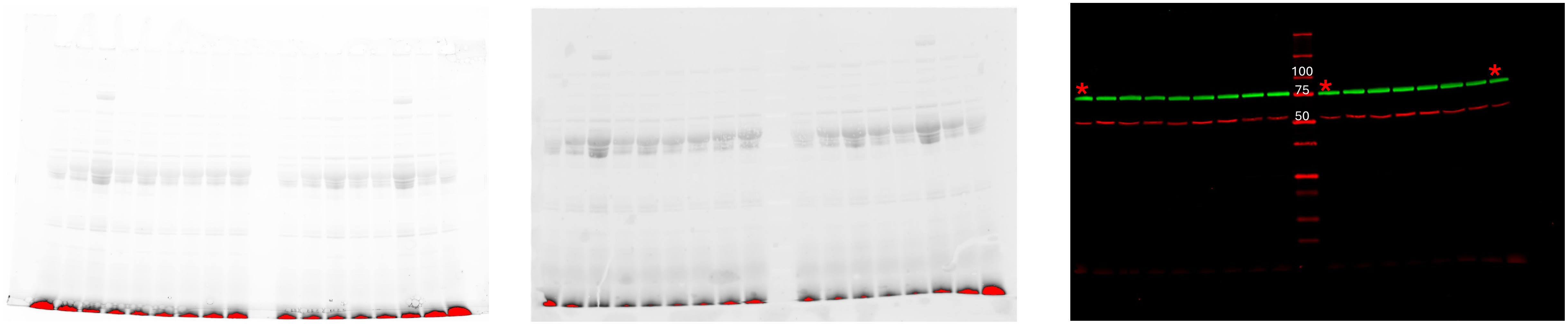


4

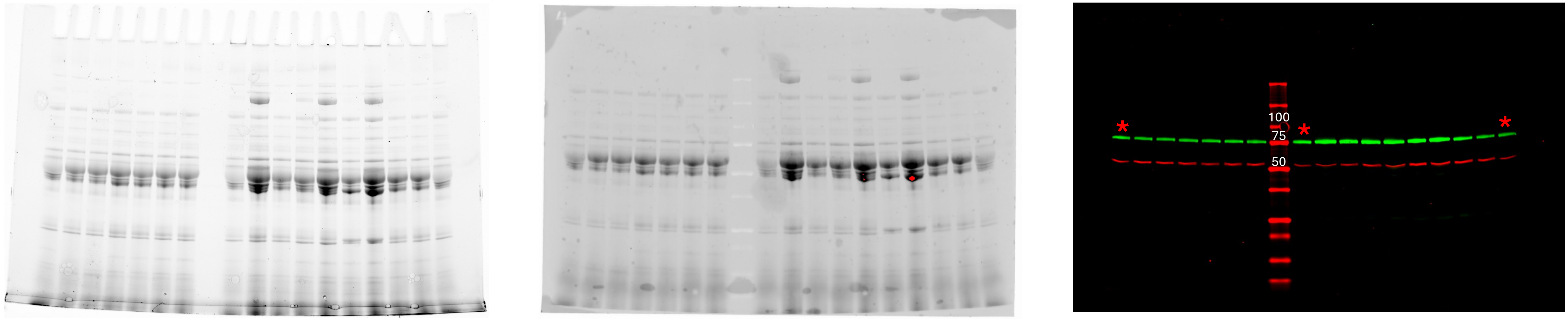

5 Gel Membrane Ab staining

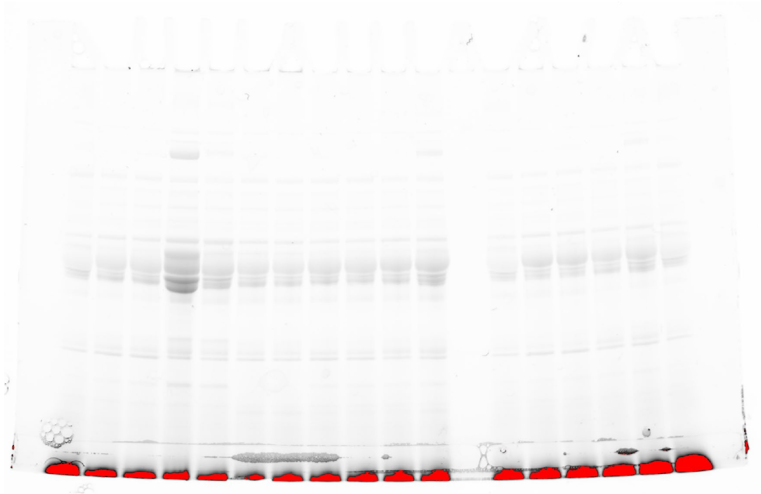

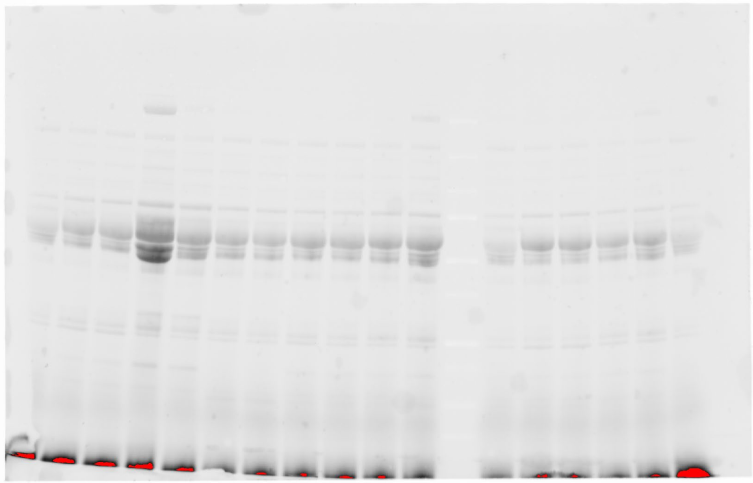

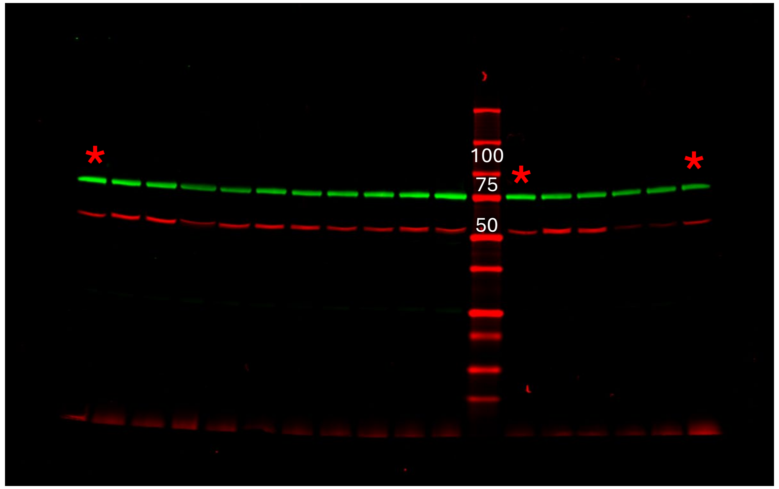


6

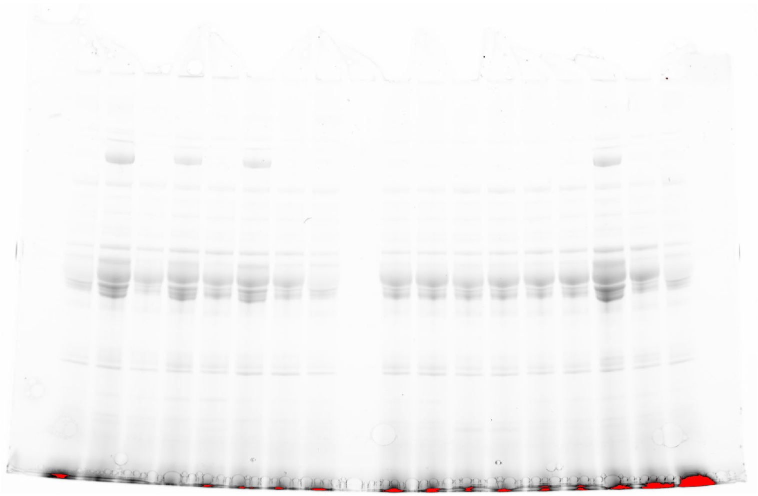

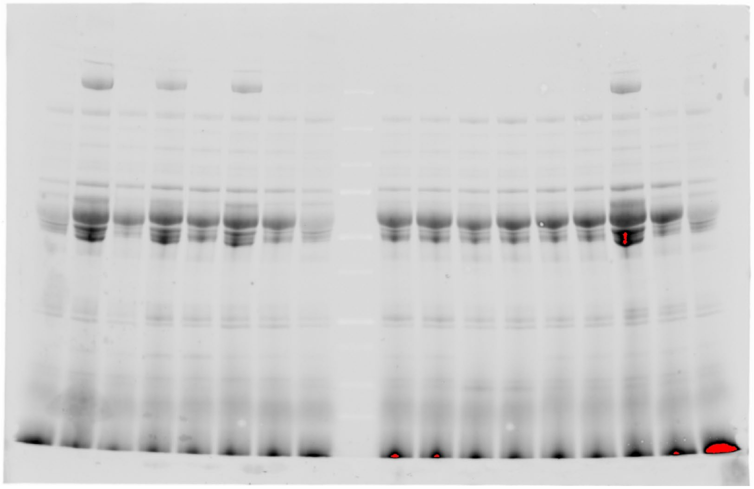

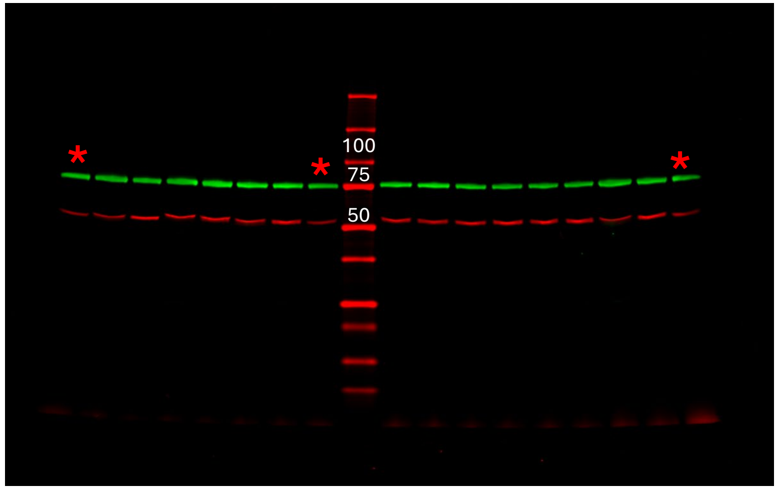

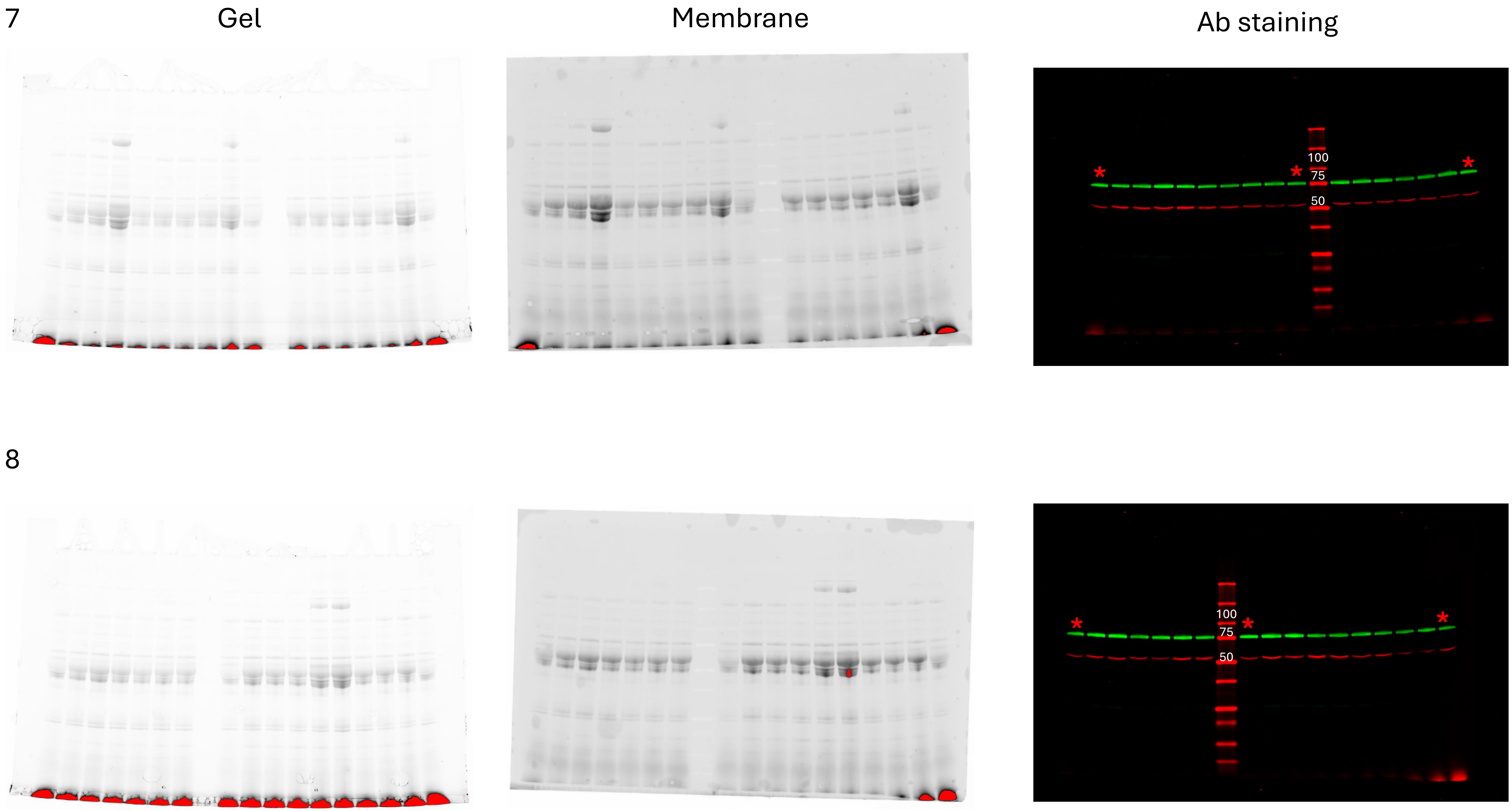
7
Gel
Membrane
Ab staining
100
75
50
*
8
100
75
50
*

9 Gel Membrane Ab staining

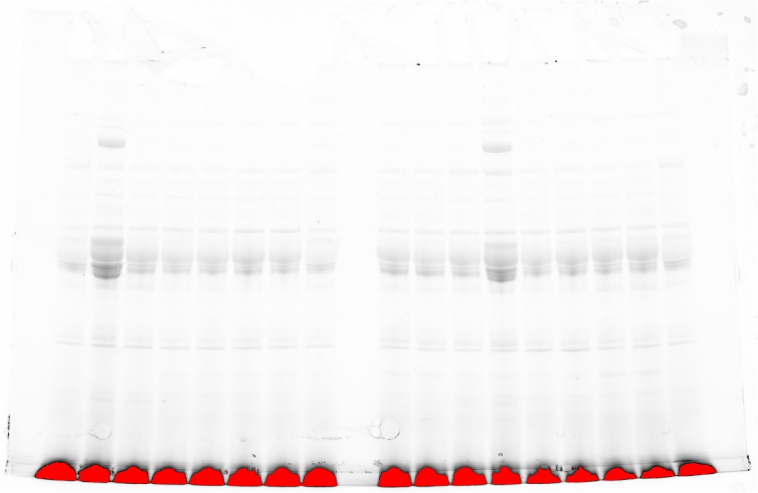

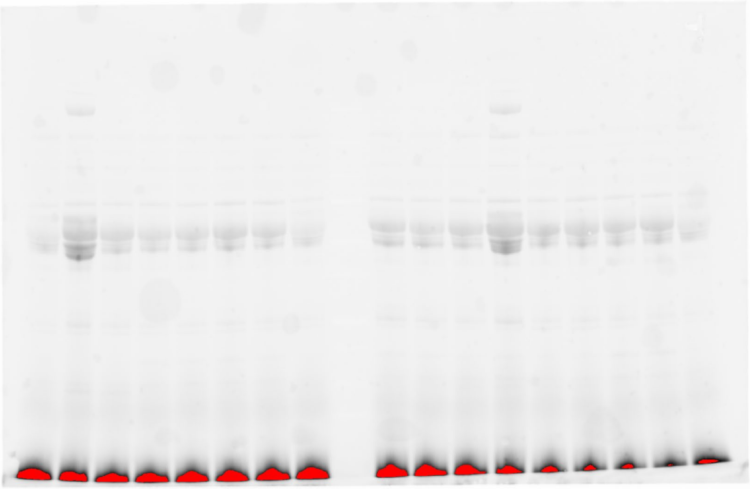

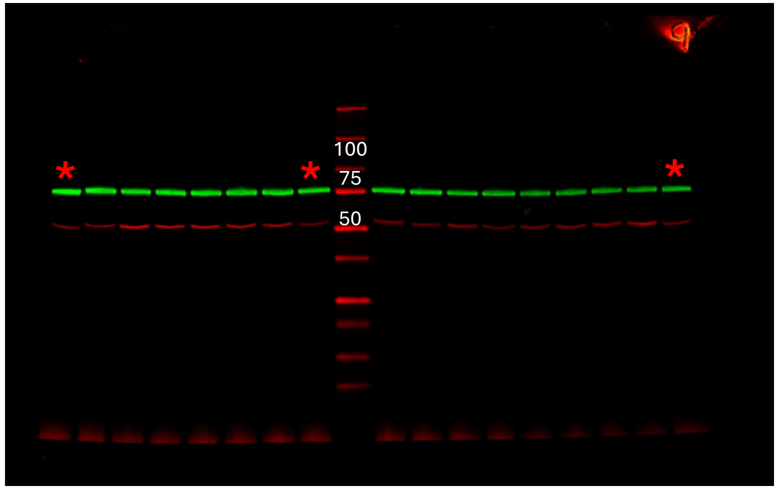


10

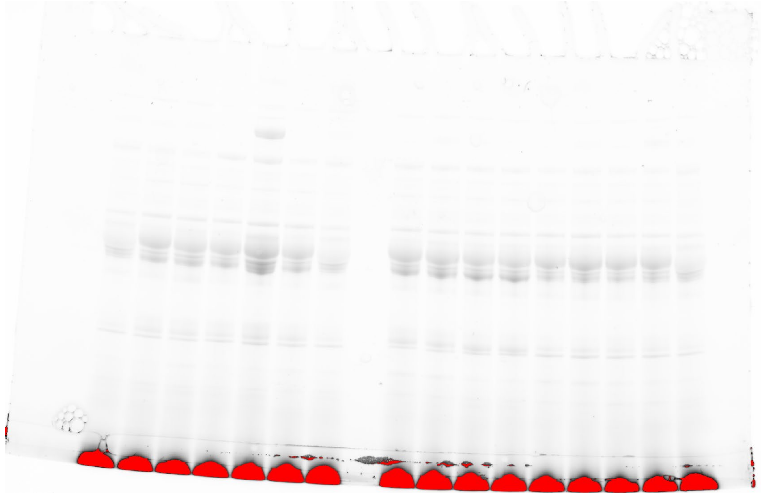

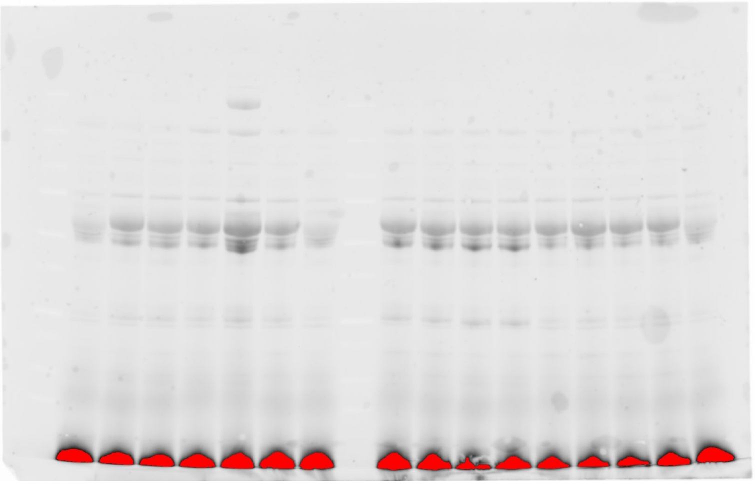

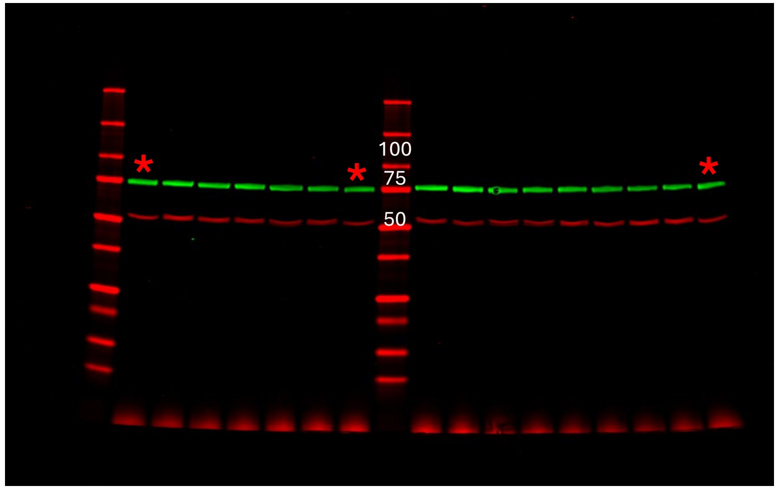

11 Gel Membrane Ab staining

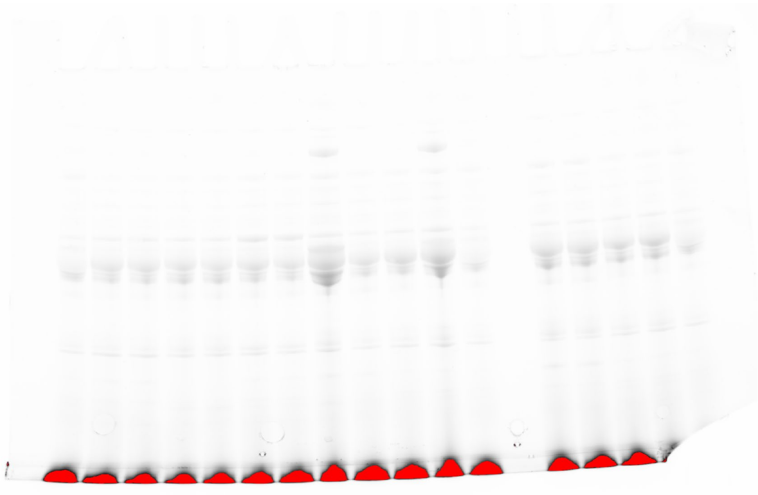

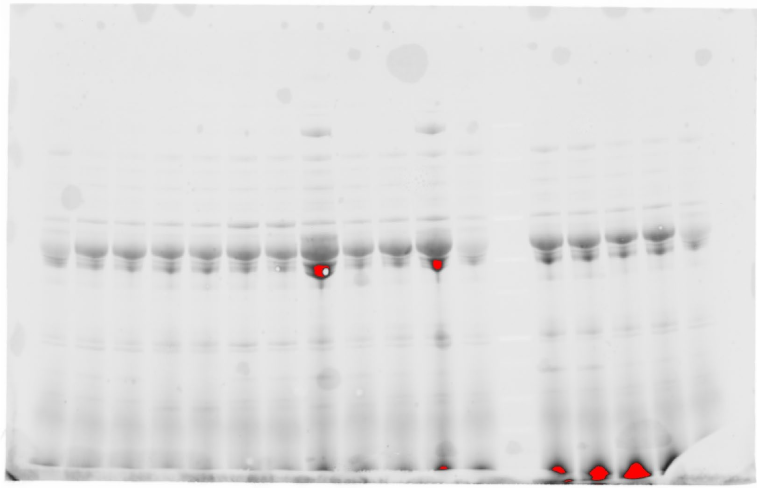

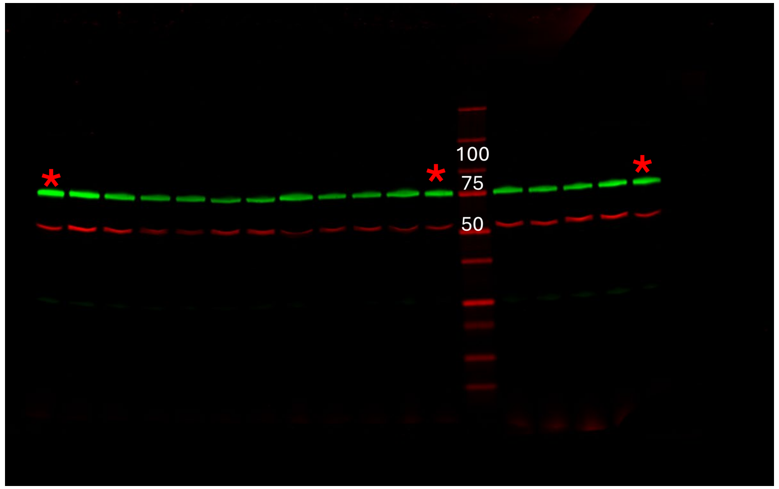


12

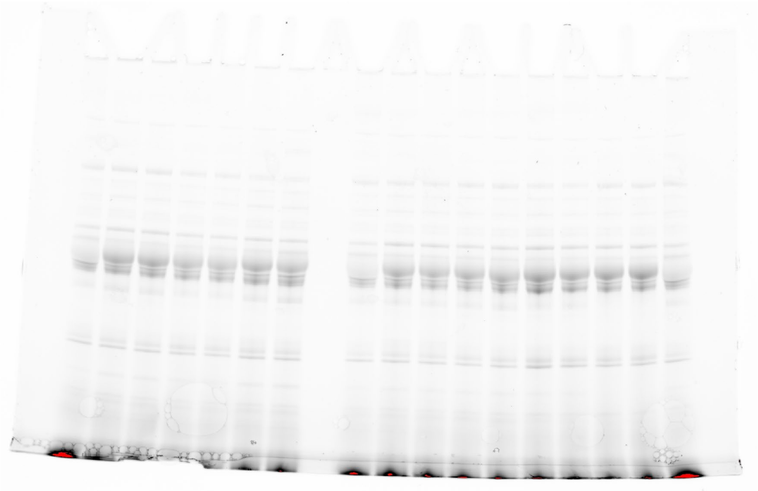

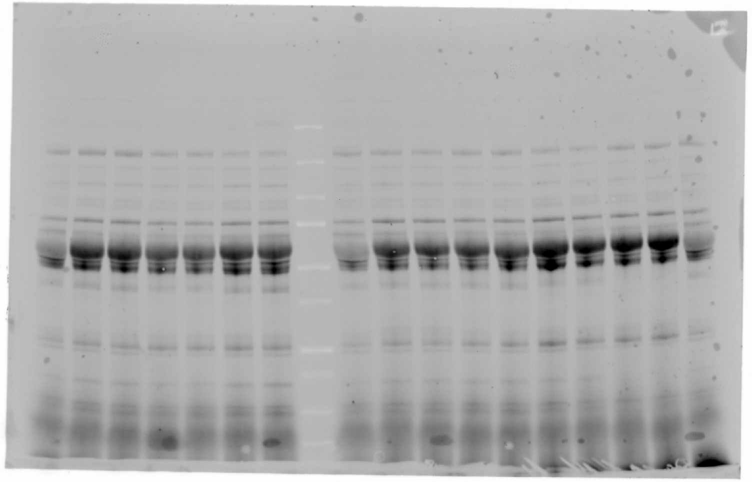

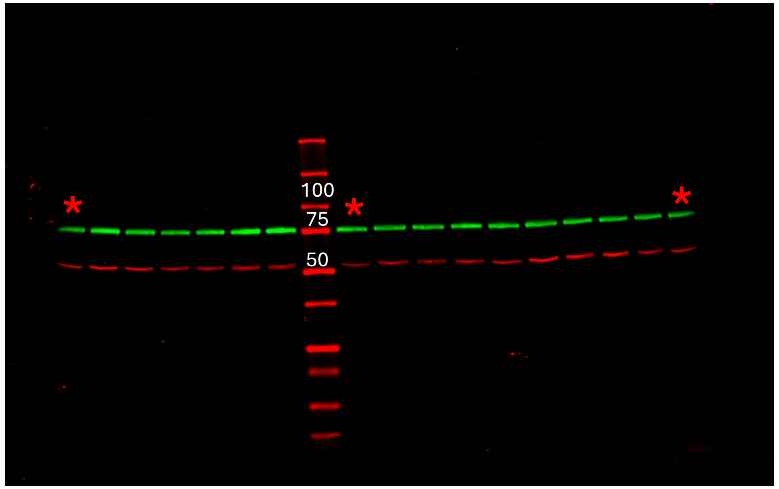

13

Gel | Membrane | Ab staining

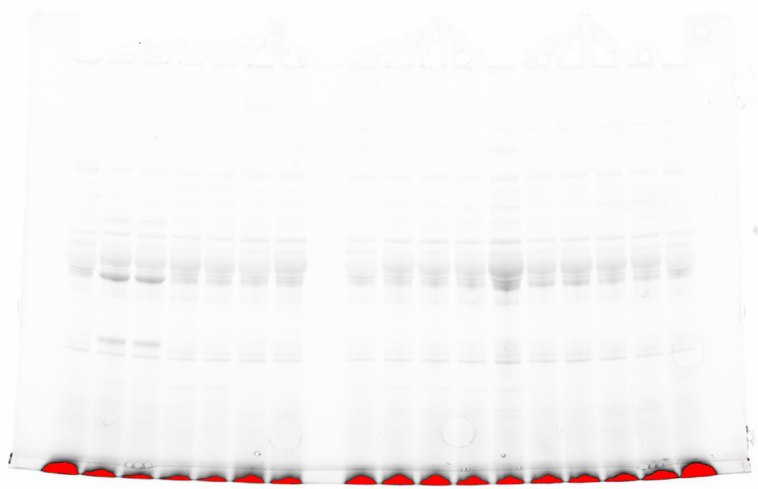

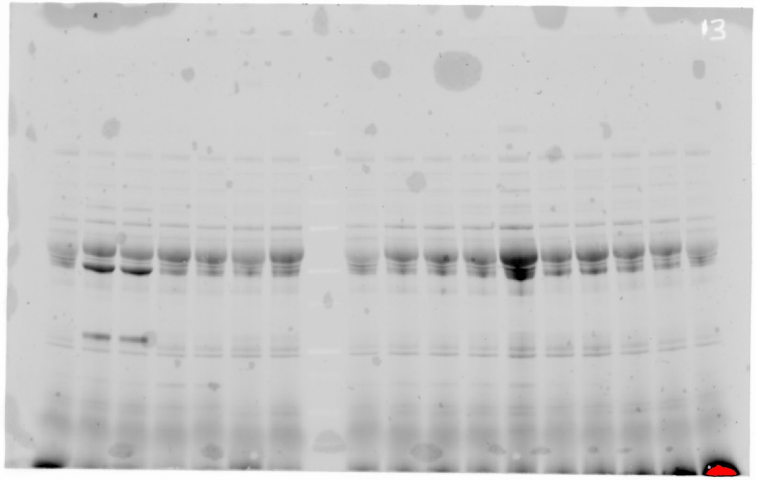


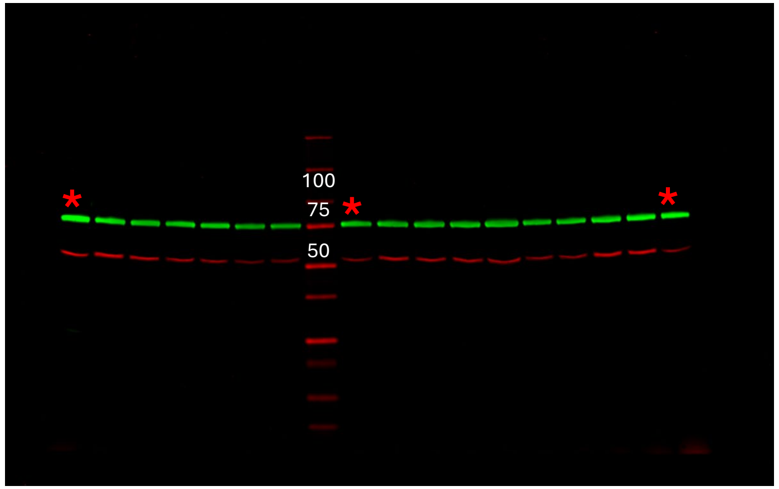


14

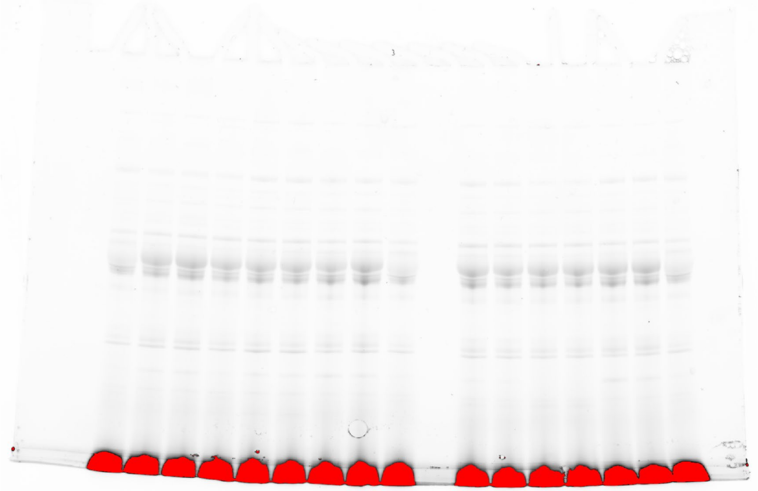

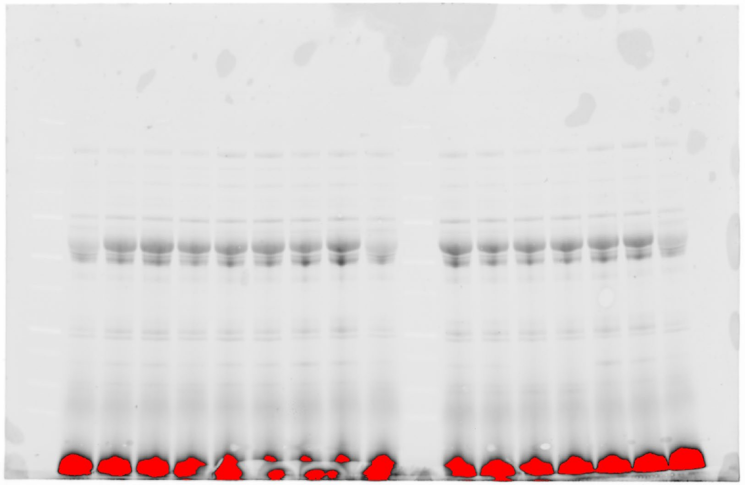

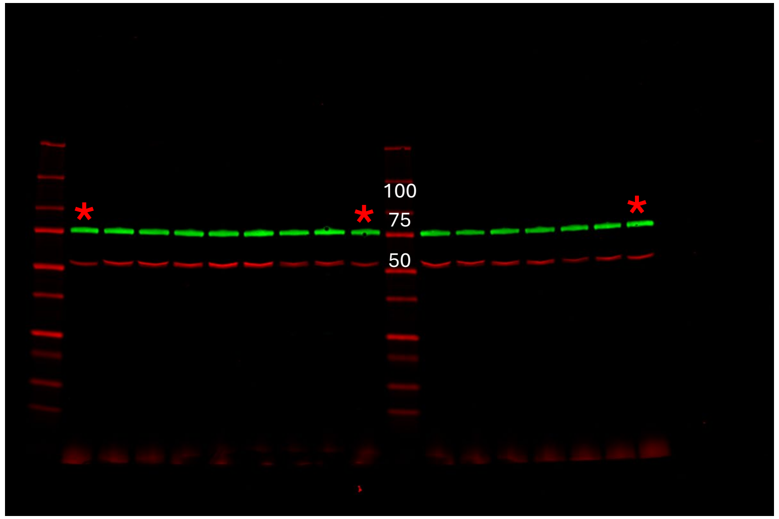

15 Gel Membrane Ab staining

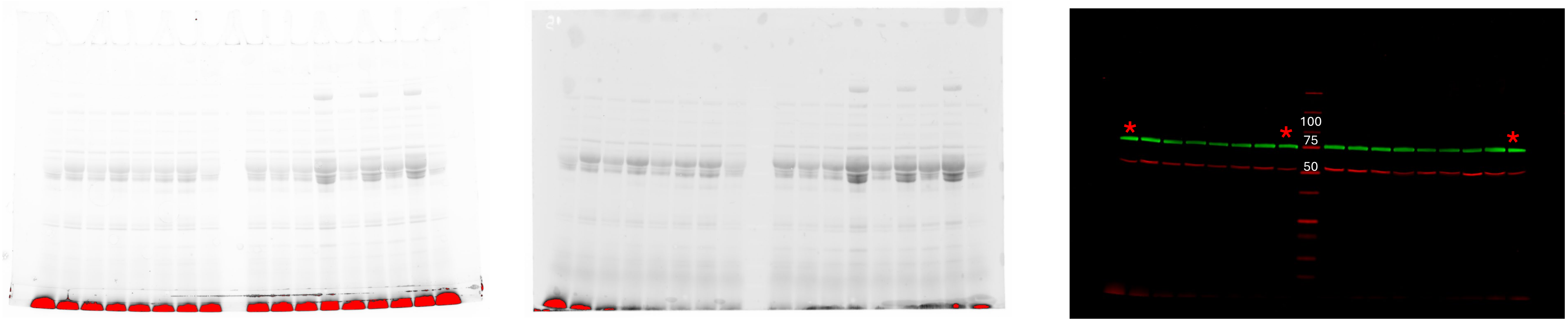


16

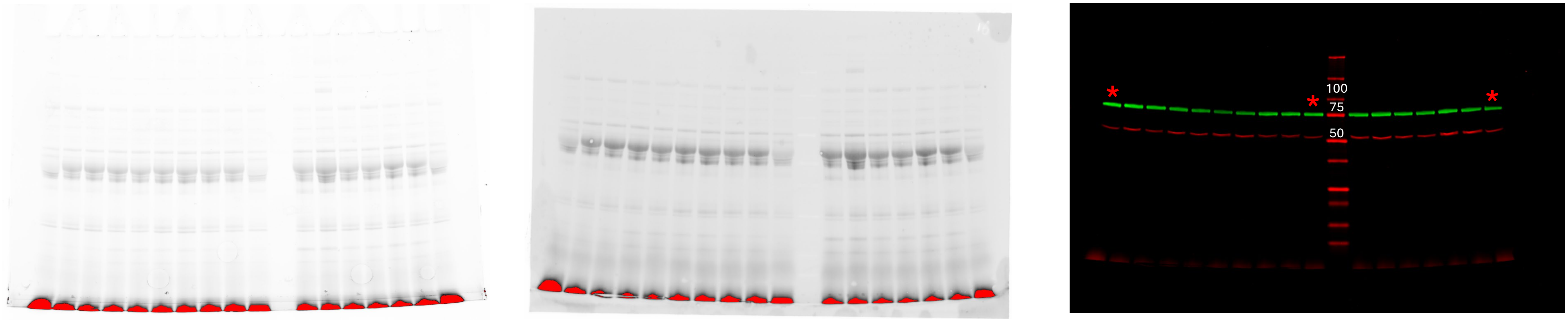

17

Gel

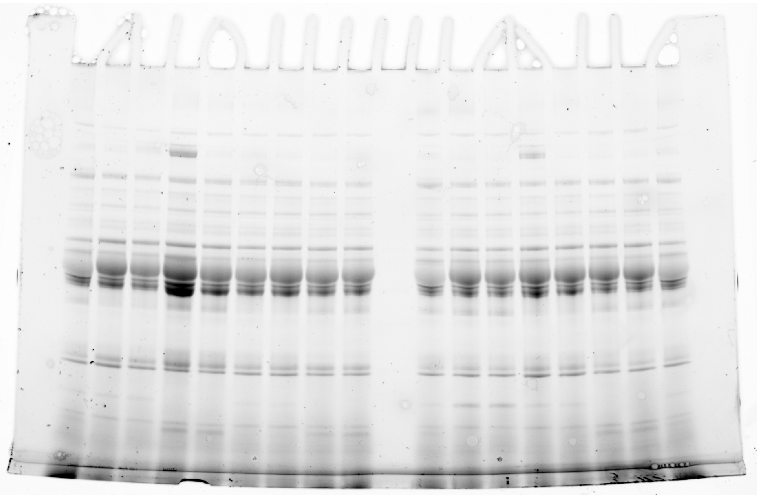

Membrane

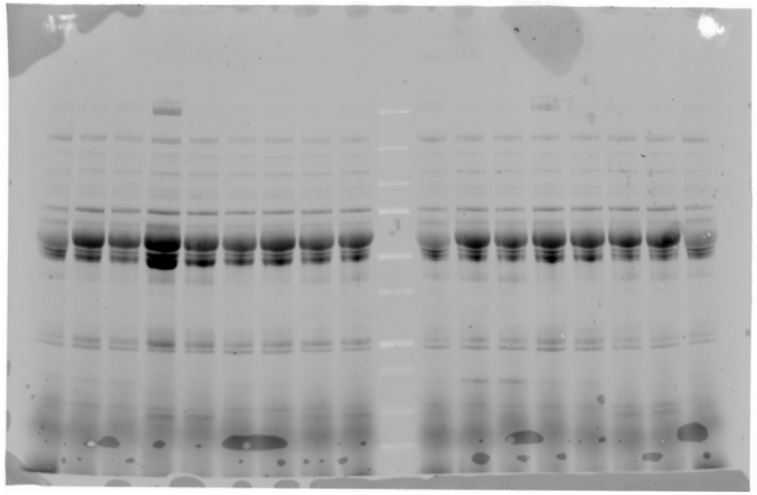

Ab staining

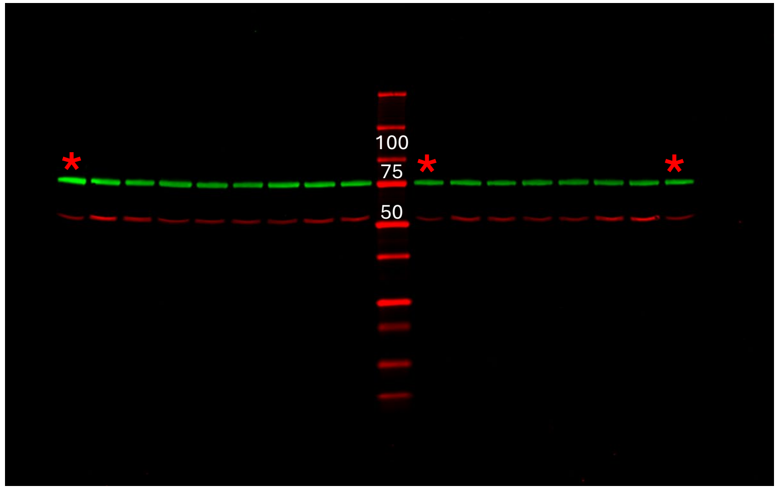
