## Supplementary figures 2 3 4 for "Circulating eNAMPT in Glaucoma: A Semi-Quantitative Plasma Analysis Before and After Nicotinamide Supplementation"

**Supplementary Figure 2.** eNAMPT/Transferrin raw signals ratio of the reference sample depending on the position in the western blot assay. eNAMPT: extracellular nicotinamide phosphoribosyltransferase.

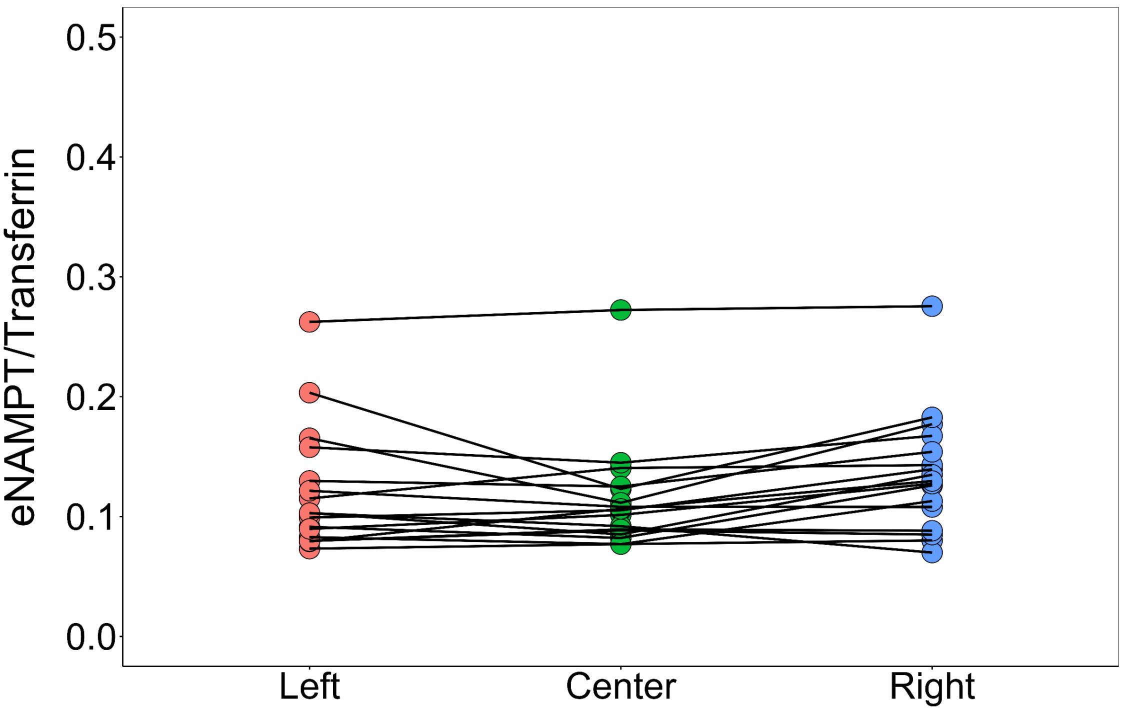

**Supplementary figure 3.** Heatmap representation of normalized relative extracellular nicotinamide phosphoribosyltransferase (eNAMPT) levels in plasma and their correlation with retinal vasculature parameters measured by optical coherence tomography angiography (OCTA) at baseline (pre-NAM treatment). Pearson correlation analyses were performed between plasma eNAMPT levels and regional OCTA perfusion metrics (FDR-adjusted p-value < 0.05 (*)). Pearson correlation coefficients are displayed within the tiles. No significant correlations were observed between eNAMPT levels and any of the measured vascular parameters. HTG: high-tension glaucoma; NTG: normal-tension glaucoma; ONH: optic nerve head; PEXG: pseudoexfoliative glaucoma.

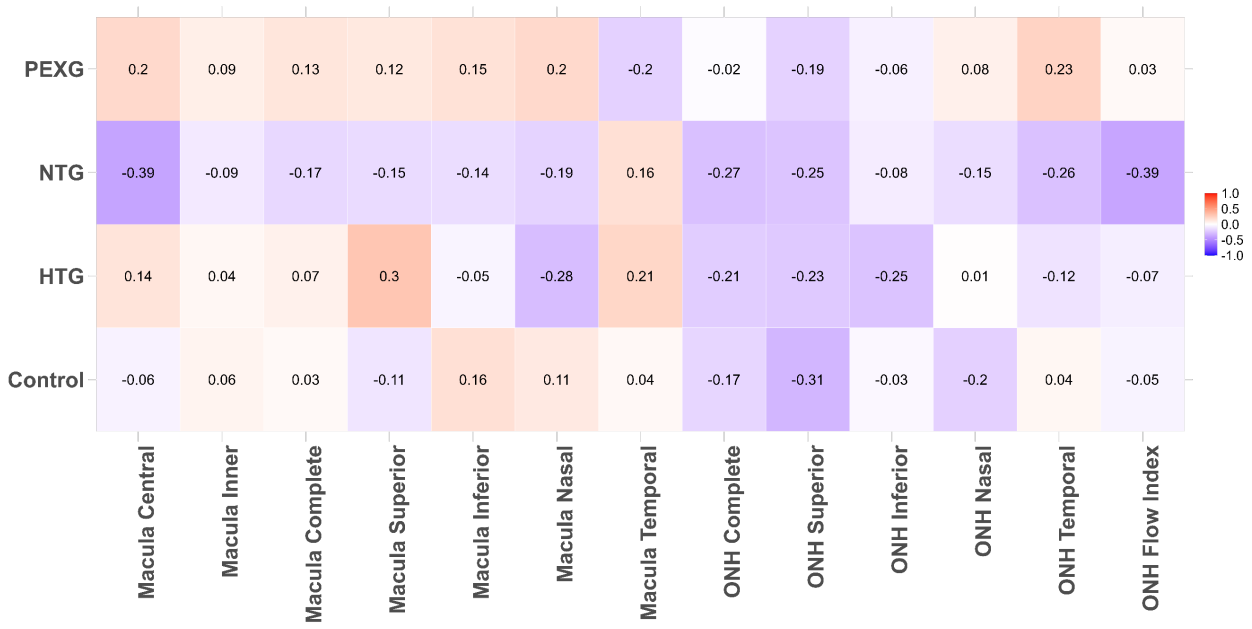

**Supplementary figure 4.** Heatmap representation of normalized relative extracellular nicotinamide phosphoribosyltransferase (eNAMPT) levels in plasma and their correlation with retinal vasculature parameters measured by optical coherence tomography angiography (OCTA) post-NAM treatment. Pearson correlation analyses were performed between plasma eNAMPT levels and regional OCTA perfusion metrics (FDR-adjusted p-value < 0.05 (*)). Pearson correlation coefficients are displayed within the tiles. No significant correlations were observed between eNAMPT levels and any of the measured vascular parameters. HTG: high-tension glaucoma; NTG: normal-tension glaucoma; ONH: optic nerve head; PEXG: pseudoexfoliative glaucoma.

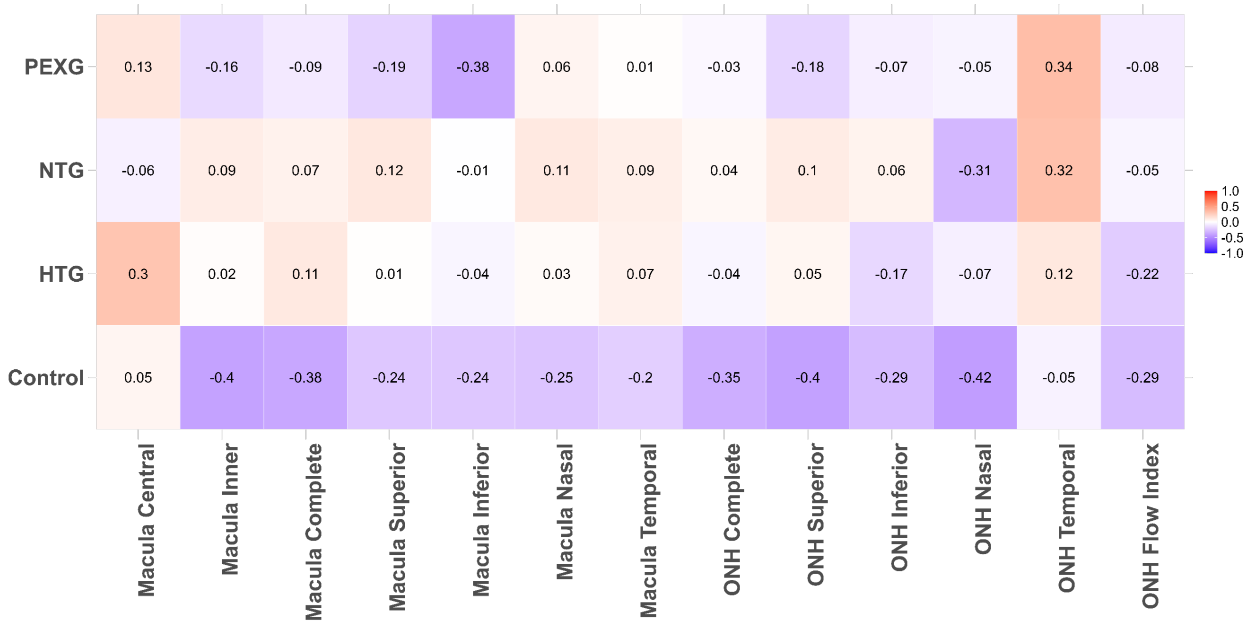
