## Supplementary tables for "Circulating eNAMPT in Glaucoma: A Semi-Quantitative Plasma Analysis Before and After Nicotinamide Supplementation"

**Supplementary Table 1.** Intra and inter-assay variability table. In the average raw signal columns and the average ratio column, the average from the respective reference sample triplicate in each blot for the band signals of extracellular nicotinamide phosphoribosyltransferase (eNAMPT), transferrin, and the ratio of their signals can be found. In the coefficient of variability (CV%) columns the CV% of extracellular nicotinamide phosphoribosyltransferase (eNAMPT) and transferrin, as well as the ratio of their signals for the reference sample is calculated within each blot. In bold the intra-assay CV% average between all blots can be found (14.9%) as well as the inter-assay CV% (37.9%).

| | | **eNAMPT** | | **Transferrin** | | **eNAMPT/Transferrin** | |
|---|---|---|---|---|---|---|---|
| | | **Average raw signal** | **CV%** | **Average raw signal** | **CV%** | **Average ratio** | **CV%** |
| **Blot 1** | | 13062 | 9.1 | 163115 | 6.3 | 0.08 | 3.6 |
| **Blot 2** | | 9171 | 4.2 | 87802 | 19.4 | 0.11 | 23.5 |
| **Blot 3** | | 18866 | 16.3 | 167694 | 15.9 | 0.11 | 6.9 |
| **Blot 4** | | 11862 | 16.5 | 71788 | 20.0 | 0.17 | 24.6 |
| **Blot 5** | | 11832 | 14.4 | 107240 | 29.9 | 0.11 | 18.9 |
| **Blot 6** | | 12395 | 7.4 | 93895 | 9.8 | 0.13 | 11.6 |
| **Blot 7** | | 10119 | 2.7 | 120031 | 3.7 | 0.08 | 5.2 |
| **Blot 8** | | 8621 | 15.2 | 90157 | 32.5 | 0.10 | 23.1 |
| **Blot 9** | | 16310 | 5.6 | 157852 | 22.6 | 0.11 | 18.2 |
| **Blot 10** | | 14458 | 13.0 | 92308 | 11.4 | 0.16 | 7.2 |
| **Blot 11** | | 34230 | 22.3 | 252216 | 23.4 | 0.14 | 11.4 |
| **Blot 12** | | 8732 | 1.0 | 60050 | 25.4 | 0.15 | 23.1 |
| **Blot 13** | | 25056 | 39.9 | 279604 | 25.1 | 0.09 | 19.0 |
| **Blot 14** | | 34917 | 6.7 | 129251 | 5.3 | 0.27 | 2.5 |
| **Blot 15** | | 15015 | 5.5 | 175468 | 6.5 | 0.09 | 6.2 |
| **Blot 16** | | 13850 | 4.8 | 163320 | 20.6 | 0.09 | 25.0 |
| **Blot 17** | | 12128 | 13.8 | 121146 | 33.9 | 0.11 | 24.0 |
| | **Average** | 15919 | 11.7 | 137231 | 18.3 | 0.12 | **14.9** |
| | **SD** | ± 8093 | ± 9.4 | ± 60193 | ± 9.7 | ± 0.05 | ± 8.4 |
| | **Min** | 8621 | 1.0 | 60050 | 3.7 | 0.08 | 2.5 |
| | **Max** | 34917 | 39.9 | 279604 | 33.9 | 0.27 | 25.0 |
| | **CV% (inter-assay)** | 50.8 | | 43.9 | | **37.9** | |

**Supplementary Table 2.** Generalized linear mixed effects model (Gamma distribution with a log link and Gaussian random effects) including age and sex as co-variates. eNAMPT: extracellular nicotinamide phosphoribosyltransferase; HTG: high tension glaucoma; N Subject: number of levels of groups in the random effects, meaning number of unique participants included in the model; NAM: nicotinamide; NTG: normal tension glaucoma; PEXG: pseudoexfoliative glaucoma; Std: standard; τ Subject: variance of the random set intercepts associated with subject, representing variability within paired participants; $\sigma^2$: residual variance; (:) indicates interaction between two variables.

| | **Normalized relative eNAMPT levels** | | |
|---|---|---|---|
| *Predictors* | *Estimate* | *Std. error* | *P-value* |
| Controls (Intercept) | 0.12730 | 0.51163 | 0.804 |
| HTG | -0.17101 | 0.11632 | 0.142 |
| NTG | -0.11845 | 0.11648 | 0.309 |
| PEXG | -0.01916 | 0.11635 | 0.869 |
| NAM Treatment | -0.02334 | 0.03212 | 0.467 |
| Sex (Male) | 0.23485 | 0.68110 | 0.730 |
| Age | 0.08061 | 0.08064 | 0.317 |
| HTG : NAM Treatment | 0.01982 | 0.04586 | 0.666 |
| NTG : NAM Treatment | -0.01136 | 0.04592 | 0.805 |
| PEXG : NAM Treatment | -0.08570 | 0.04618 | 0.063 |
| **Random Effects** | | | |
| $\sigma^2$ | 0.02841 | | |
| $\tau_{Subject}$ | 0.06 | | |
| $N_{Subject}$ | 120 | | |
| Observations | 236 | | |
